# Dominant Clade-featured SARS-CoV-2 Co-occurring Mutations Reveals Plausible Epistasis: An *in silico* based Hypothetical Model

**DOI:** 10.1101/2021.02.21.21252137

**Authors:** A. S. M. Rubayet Ul Alam, Ovinu Kibria Islam, Md. Shazid Hasan, Mir Raihanul Islam, Shafi Mahmud, Hassan M. Al□Emran, Iqbal Kabir Jahid, Keith A. Crandall, M. Anwar Hossain

**Affiliations:** Department of Microbiology, Jashore University of Science and Technology, Jashore-7408, Bangladesh; BRAC James P Grant School of Public Health, BRAC University, Bangladesh; Genetic Engineering and Biotechnology, University of Rajshahi, Rajshai-6205, Bangladesh; Department of Biomedical Engineering, Jashore University of Science and Technology, Jashore-7408, Bangladesh; Computational Biology Institute and Department of Biostatistics & Bioinformatics, Milken Institute School of Public Health, The George Washington University, Washington, DC, USA; Jashore University of Science and Technology, Jashore-7408, Bangladesh; Department of Microbiology, University of Dhaka, Dhaka-1000, Bangladesh

**Keywords:** SARS-CoV-2, COVID-19, Infection Paradox, Fitness, Virulence, Clades, Co-occurring mutations

## Abstract

SARS-CoV-2 is evolved into eight fundamental clades where four (G, GH, GR, and GV) are globally prevalent in 2020. How the featured co-occurring mutations of these clades are linked with viral fitness is the main question here and we thus proposed a hypothetical model using *in silico* approach to explain the plausible epistatic effects of those mutations on viral replication and transmission. Molecular docking and dynamics analyses showed the higher infectiousness of a spike mutant through more favorable binding of G_614_ with the elastase-2. RdRp mutation p.P323L significantly increased genome-wide mutations (p<0.0001) since more flexible RdRp (mutated)-NSP8 interaction may accelerate replication. Superior RNA stability and structural variation at NSP3:C241T might impact protein and/or RNA interactions. Another silent 5’UTR:C241T mutation might affect translational efficiency and viral packaging. These four G-clade-featured co-occurring mutations might increase viral replication. Sentinel GH-clade ORF3a:p.Q57H constricted ion-channel through inter-transmembrane-domain interaction of cysteine(C81)-histidine(H57) and GR-clade N:p.RG203-204KR would stabilize RNA interaction by a more flexible and hypo-phosphorylated SR-rich region. GV-clade viruses seemingly gained the evolutionary advantage of the confounding factors; nevertheless, N:p.A220V might modulate RNA binding with no phenotypic effect. Our hypothetical model needs further retrospective and prospective studies to understand detailed molecular events featuring the fitness of SARS-CoV-2.

## 1. Introduction

Severe acute respiratory syndrome coronavirus 2 (SARS-CoV-2), the etiological agent of COVID-19 pandemic, has gained some extraordinary attributes that make it extremely infectious: High replication rate, large burst size, high stability in the environment, strong binding efficiency of spike glycoprotein (S) receptor-binding domain (RBD) with human angiotensin-converting enzyme 2 (ACE2) receptor, and additional furin cleavage site in S protein^1-3^. In addition to those, it has proofreading capability ensuring relatively high-fidelity replication^4^. The virus contains four major structural proteins: spike glycoprotein (S), envelope (E), membrane (M), and nucleocapsid (N) protein along with 16 nonstructural proteins (NSP1 to NSP16) and seven accessory proteins (ORF3a, ORF6, ORF7a, ORF7b, ORF8a, ORF8b, and ORF10)^5,6^. Mutational spectra within the SARS-CoV-2 genome^7,8^, spike protein^9^, RdRp^10^, ORF3a^11^, and N protein^12^ were reported.

SARS-CoV-2 was classified into eight major clades, such as G, GH, GR, GV, S, V, L, and O by global initiative on sharing all influenza data (GISAID) consortium (https://www.gisaid.org/) based on the dominant core mutations in genomes where four clades (G, GH, GR, and GV) are globally and geographically prevalent in 2020^13^. Yin ^14^ reported that the 5’
sUTR mutation 241C > T is co-occurring with three other mutations, 3037C > T (NSP3: C318T), 14408C > T (RdRp: p.P323L), and 23403A > G (S: p.D614G).

GISAID referred to these co-occurring mutations containing viruses as clade G (named after the spike D614G mutation) or PANGO (https://cov-lineages.org/) lineage B.1^15,16^. The GR clade or lineage B.1.1.* is classified with additional trinucleotide mutations at 28881-28883 (GGG>AAC); creating two consecutive amino acid changes, R203K and G204R, in N protein. Another derivative of G clade is GH or lineage B.1.*, characterized by an additional ORF3a:p.Q57H mutation. The variant GV or lineage B.1.177 featured an A222V mutation in the S protein along with other mutations of the clade G^13,16^. Also, N: A220V, ORF10: V30L and three other synonymous mutations T445C, C6286T, and C26801G are observed for this clade^17^.

The most frequently observed mutation is D614G of the S protein^18^, which has direct roles in receptor binding, and immunogenicity, thus viral immune-escape, transmission, and replication fitness^19,20^. Mutations in proteins other than spike could also affect viral pathogenicity and transmissibility, but the role of those dominant clade-featured mutations has remained largely underestimated. Although the possible role of ORF3a:p.Q57H in replication cycle^21^ has recently been investigated, the molecular perspective was not fully explained there. The effect of 5’UTR: C241T, Leader: T445C, NSP3: C318T, RdRp:p.P323L, N:p.RG203-203KR, and N:p.A220V is still being overlooked.

Different mutation(s) of SARS-CoV-2 may work independently or through epistatic interactions^22,23^; however, it is difficult to determine exactly how these co-occurring mutations, if not all, might have gained their selective evolutionary fitness^22,24,25^. Hence, many hypothetical questions remain: What are the impacts of these mutations on protein structures, and what can be their functional roles? How might these mutated proteins interact together? Is there any possible role of the co-occurring ‘silent’ mutation? Could these mutations have any plausible impact on viral fitness and virulence? We attempt here to answer these questions by *in silico* molecular insights of SARS-CoV-2 mutants and possible interactions of proteins containing co-occurring mutations. Overall, this study aims to determine plausible individual and/or epistatic impacts of those mutants during replication in terms of viral entry and fusion, evasion of host cell lysis, replication rate, ribonucleoprotein stability, protein-protein interactions, translational capacity, and ultimately the probable combined effect on viral transmission and fitness.

## 2. Materials and Methods

### 2.1 Retrieval of Sequences and Mutation Analyses

This study analyzed 225,526 high-coverage (<1% Ns and <0.05% unique amino acid mutations) and complete (>29,000 nucleotides) genome sequences from a total of 3,16,166 sequences submitted to GISAID from January 01, 2020, to January 03, 2021. We removed the non-human host-generated sequences during dataset preparation. The Wuhan-Hu-1 (Accession ID-NC_045512.2) isolate was used as the reference genome.

A python script (https://github.com/hridoy04/counting-mutations) was used to partition a significant part of the dataset into two subsets based on the RdRp: C14408T mutation and estimated the genome-wide variations (single nucleotide changes) for each strain. For the genome-wide mutation analysis, a total of 37,179 sequences (RdRp wild type or ‘C’ variant: 9,815; and mutant or ‘T’ variant: 27,364) were analyzed from our dataset. The frequency of mutations was tested for significance with the Wilcoxon signed-rank test between RdRp ‘C’ variant and ‘T’ variant using IBM SPSS statistics 25.

### 2.2 Stability, Secondary and Three-Dimensional Structure Prediction Analyses of S, RdRp, ORF3a, and N Proteins

DynaMut^26^ and FoldX 5.0^27,28^ were used to determine the stability of both wild and mutant variants of N, RdRp, S, and ORF3a proteins. PredictProtein^29^ was utilized for analyzing and predicting the possible secondary structure and solvent accessibility of both wild and mutant variants of those proteins. The SWISS-MODEL homology modeling webtool^30^ was utilized for generating the three-dimensional (3D) structures of the RdRp, S, and ORF3a protein using 7c2k.1.A, 6xr8.1.A, and 6xdc.1.A PDB structure as the template, respectively. Modeller v9.25^31^ was also used to generate the structures against the same templates. I-TASSER^32^ with default protein modeling mode was employed to construct the N protein 3D structure of wild and mutant type since there was no template structure available for the protein. The built-in structural assessment tools (Ramachandran plot, MolProbity, and Quality estimate) of SWISS-MODEL were used to check the quality of generated structures.

### 2.3 Molecular Docking and Dynamics of RdRp-NSP8 and Spike-Elastase2 Complexes

Determination of the active sites affected by binding is a prerequisite for docking analysis. We chose 323 along with the surrounding residues (315-324) of RdRp and the residues 110 to 122 of NSP8 monomer as the active sites based on the previously reported structure^33^. The passive residues were defined automatically where all surface residues were selected within the 6.5°A radius around the active residues. The molecular docking of the wild and predicted mutated RdRp with the NSP8 monomer from the PDB structure 7C2K was performed using the HADDOCKv2.4 to evaluate the interaction^34^. The binding affinity of the docked RdRp-NSP8 complex was predicted using the PRODIGY^35^. The number and specific interfacial contacts (IC) for each of the complexes were identified.

The human neutrophil elastase (hNE) or elastase-2 (PDB id: 5A0C) was chosen for docking of the S protein, based on earlier reports^36^. Here we employed CPORT^37^ to find out the active and passive protein-protein interface residues of hNE. The S protein active sites were chosen based on the target region (594-638) interacting with the elastase-2. The passive residues of S protein were defined automatically as mentioned for RdRp-NSP8 docking analysis. Afterward, we individually docked wild (614D) and mutated (614G) S protein with the hNE using HADDOCK 2.4. The binding affinity of the docked complexes, as well as the number and specific interfacial contacts (IC), were predicted as performed after RdRP-NSP8 docking. We employed HDOCK server^38^ with specifying the active binding sites residues for predicting the molecular docking energy.

The structural stability of the above-mentioned protein complexes (RdRP-NSP8 and Spike-Elastase2) and their variations were assessed through YASARA Dynamics software package (Land & Humble, 2018). We used AMBER14 force field (Dickson et al., 2014) for these four systems, and the cubic simulation cell was created with the TIP3P (at 0.997 g/L-1, 25C, and 1 atm) water solvation model. The PME or particle mesh Ewald methods were applied to calculate the long-range electrostatic interaction by a cut-off radius of 8Å^39^. We applied the Berendsen thermostat to maintain the temperature of the simulation cell. The time step of the simulation was set as 1.25fs^40^ and the simulation trajectories were saved after every 100ps. Finally, we conducted the molecular dynamics simulation for 100ns^41-44^.

### 2.4 Mutational Analysis of Transmembrane Domain 1 of ORF3a and serine-rich (SR) domain of N protein

The complete genome of 12 pangolin-derived coronavirus strains, as well as 38 bats, civet, and human SARS-CoVs, were downloaded from GISAID and NCBI, respectively for the mutational comparison between the SARS-CoV and SARS-CoV-2. We mainly targeted transmembrane domain 1 (TM1), which covers 41 to 63 residues, of ORF3a to find the identical mutation and scan overall variation in TM1. A generalized comparison between SARS-CoV and SARS-CoV-2 reference sequences was performed to identify the mutations in the SR-rich region that will help to postulate on N protein functions of novel coronavirus based on previous related research on SARS-CoV.

### 2.5 Analyzing RNA Folding prediction of 5’ sUTR, Leader protein, and NSP3

The Mfold web server^45^ was used with default parameters to check the folding pattern of RNA secondary structure in the mutated 5’
sUTR, synonymous leader (T445C), and NSP3 region (C3037T). The change C6286T is in between the nucleic acid-binding (NAB) domain and betacoronavirus specific marker (βSM) domain of the NSP3 region. The change C26801G is at the transmembrane region 3 (TM3) of the virion membrane. Thus, changing in C6286T and C26801G will not affect the function significantly and was not predicted here. The structure of complete mutant 5’UTR (variant ‘T’) was compared with the wild type (variant ‘C’) secondary pattern as mentioned in the Huston et al. (2021) ^46^. From the Mfold web server, we also estimated free energy change (∆G) for wild and mutant leader and NSP3 RNA fold to find any variation in stability.

## 3. RESULTS AND DISCUSSION

The possible individual effects of a total nine mutations in S, RdRp, ORF3a, N, 5’UTR, leader protein (NSP1), and NSP3 in viral replication cycle and transmission were discussed with associated results. Zeng et al. (2020) showed the links of these mutations towards possible epistatic effects on fitness using statistical analysis that duly suits our purpose of presenting how the mutations might play the combined roles^23^. Whereas researches on molecular docking of the spike protein^47,48^ and RdRp^49,50^ in search of potential drug targets is a continuous process, our study approached in a unique way to dock spike with elastase-2 and RdRp with NSP8 to satisfy our purposes. The overall epistatic interactions of the mutant proteins and/or RNA was then depicted (Figure 1) as a hypothetical model and discussed.

**Figure 1.**
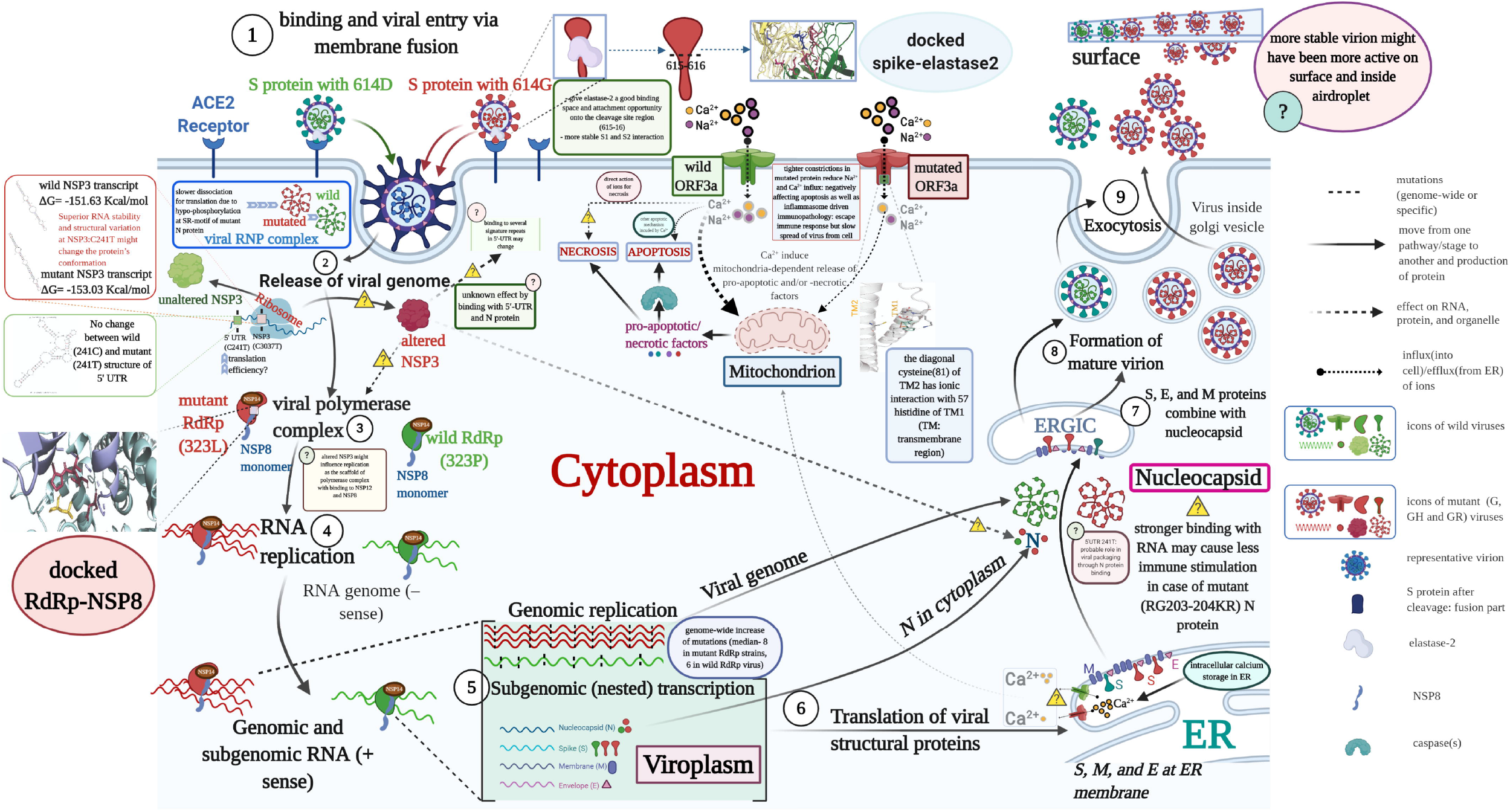
Schematic diagram of SARS-CoV-2 replication in cell showcasing the related to S, N, ORF3a, RdRp, NSP3 and 5’-UTR based epistatic interactions. The replication cycle starts with the ACE2 receptor binding of the spike glycoprotein (S) as cornered at the top-left and finishes with the exocytosis at the top-right. The viruses which do not carry G-, GH- and/or GR-featured mutations in the S, N, ORF3a, RdRp, NSP3 and 5’- UTR are denoted as the wild type where mutants contain those. Throughout the diagram, the red and green color icons such as proteins, genome, and virion represent the wild and mutant type, respectively. For a generalized virion, we used the blue color. Although this theme is not show the co-infection of both types, which might occur in rare occasions, we showed the comparative epistatic effects side-by-side fashion during the whole replication cycle that will make it easy to grasp. Related figure(s) for each protein are shown in the enclosed box. To mean the uncertainty or unknown effects of any mutant proteins/RNA structure, we used the ‘question’ mark in a pathway and on explanatory box. RdRp-RNA dependent RNA polymerase; NSP14-proof-reading enzyme of SARS-CoV-2; ER-endoplasmic reticulum; ERGIC-endoplasmic

### 3.1 Spike Protein D614G Mutation Favors Elastase-2 Binding

This study found interesting structural features of the S protein while comparing and superimposing the wild protein (D_614_) over mutated protein (G_614_). The secondary structure prediction and surface accessibility analyses showed that there was a slight mismatch at the S1-S2 junction (^681^<u>PRRA</u>R↓S^686^) where serine at 686 (S^686^) was found covered in G_614_ and exposed to the surface in D_614_. However, S^686^ in both G_614_ and D_614_ were exposed to an open-loop region to have possible contact with the proteases (supplementary Figure S1). Further investigation on the aligned 3D structures however showed no conformational change at the S1-S2 cleavage site (Figure 2c). We also observed no structural variation in the surrounding residues of the protease-targeting S1-S2 site (Figure 2c), which eliminated the assumption of Phan^51^. The predictive 3D models and structural assessment of D_614_ and G_614_ variants confirmed that the cleavage site at 815-16 of S2 subunit (^812^PSKR↓S^816^) or S2’^1,52^ had no structural and surface topological variation (Figure 2d-e). Rather, the superimposed 3D structures suggested a conformational change in the immediate downstream region (^618^TEVPVAIHADQLTPT^632^) of the 614^th^ position of mutated protein (G_614_) that was not observed in D_614_ variants (Figure 2a-b).

**Figure 2.**
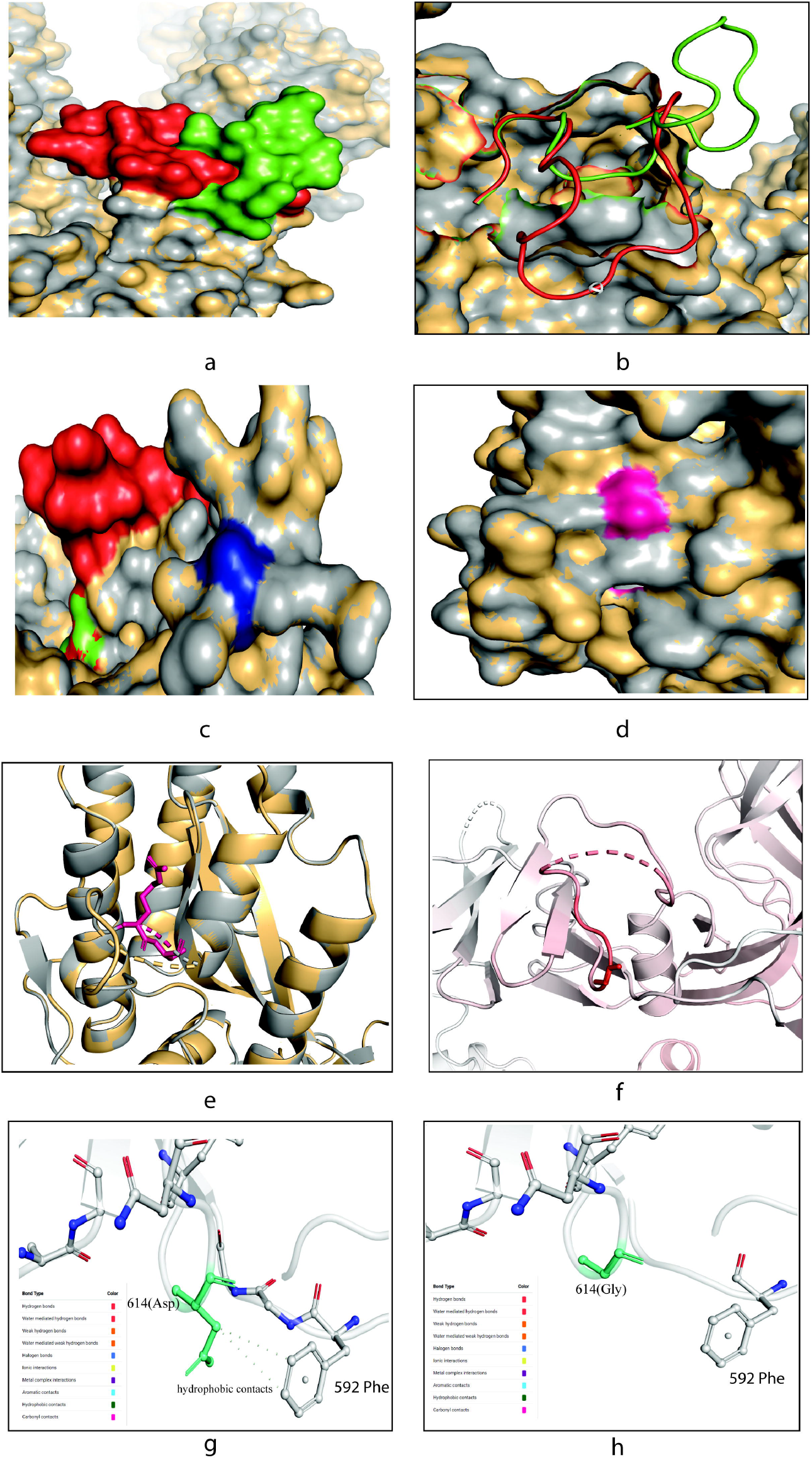
Different structural and stability comparison of wild and mutant spike protein. Structural superposition of wild and mutant spike proteins **(a-b)**; conformation in the S1-S2 **(c)** and S2’sites **(d-e)**; representation of vibrational entropy energy change on the mutant type structure **(f)**; and interatomic interaction prediction of both wild **(g)** and mutant **(h)** types. For Figure a-e, the gray and yellow color represent the wild and mutant protein, respectively. **(a)** The downstream (617-636) of D614G in wild (green) and mutant (red) S protein was focused. Overlapping of the wild (D_614_) and mutant (G_614_) S protein showed conformational change in the 3D structures. **(b)** However, the conformational change are in the loop region (618-632) of the proteins thus may potentially play role in interacting with other proteins or enzymes, such as elastase-2 as we focused in this work. **(c)** No change was found in the S1-S2 cleavage site (685-686), depicted in blue color, of the wild and mutant protein. **(d)** Surface and **(e)** cartoon (2°) structure of the superimposed wild and mutant proteins where the S2’ (pink) is situated in surface region and do not show any change in accessibility in the residual loop region. **(f)** The mutant (G_614_) protein showed higher flexibility in the G^614^ (sticks) and its surroundings (red). The intra-molecular interaction determined the overall stability of the **(g)** wild and **(h)** mutant structure where C_β_ of D^614^ (aspartic acid at 614; green stick modelled) had two hydrophobic interaction with the benzene rings. This intramolecular contacts stabilize the S protein of wild type and missing of this bond destabilize the mutant (G_614_) protein. The mutant protein has glycine at 614 which has less chance of interacting with other neighboring amino acids due to its shorter and nonpolar R-group. The color code representing the bond type is presented in each **(g)** and **(h)**.

Several experiments suggested that mutated (G_614_) protein contains a novel serine protease cleavage site at 615-616 that is cleaved by host elastase-2, a potent neutrophil elastase, level of which at the site of infection during inflammation will facilitate the host cell entry for G_614_^18,36,53,54^. The elastase-2 restrictedly cut valine at 615, due to its valine-dependent constriction of catalytic groove^55^. The present sequence setting surrounding G_614_ (P6-^610^VLYQGV↓NCTEV^620^-P’5) showed a higher enzymatic activity on the spike^36^, which cannot be completely aligned with the previous works on the sequence-based substrate specificity of elastase-2^56^. However, the first non-aligned residue of the superimposed G_614_, located at the P’4 position (T^618^), may also be important for binding with the elastase-2, and further down the threonine (T) at 618, the residues may affect the bonding with the respective amino acids of elastase-2 (Figure 2a-b). This changed conformation at the downstream binding site of G_614_ may help overcome unfavorable adjacent sequence motifs around G^614^ residue. Therefore, the simultaneous or sequential processing of the mutated S protein by TMPRSS2/Furin/Cathepsin and/or elastase-2 facilitates a more efficient SARS-CoV-2 entry into the host cells and cell-cell fusion^36,53,54^.

This study further observed the possible association of the S protein with elastase-2 and found an increased binding affinity in case of G_614_ (Table 1). Hence, the active sites of the mutated protein interacted efficiently with more amino acids of elastase-2 (Table 2), possibly providing a better catalytic activity as shown by Hu et al. (2020)^36^. The mutation may have changed the structural configuration of the elastase-2 cleavage site in a way that the enzyme is facing less challenge to get near to the cutting site of the altered protein (Figure 2a-b and 3). The efficient cleaving of this enzyme, although located in an upstream position of the S1-S2 junction, may assist in releasing S1 from S2 and change the conformation in a way that later help in cleaving at the S2’ site by other protease(s) before fusion^57,58^. Mutated spike protein and elastase-2 complex was more flexible than the wild spike-elastase complex, and the interactions with enzyme was also different as shown in RMSD deviation between the complexes (Figure 4).

**Table 1.**
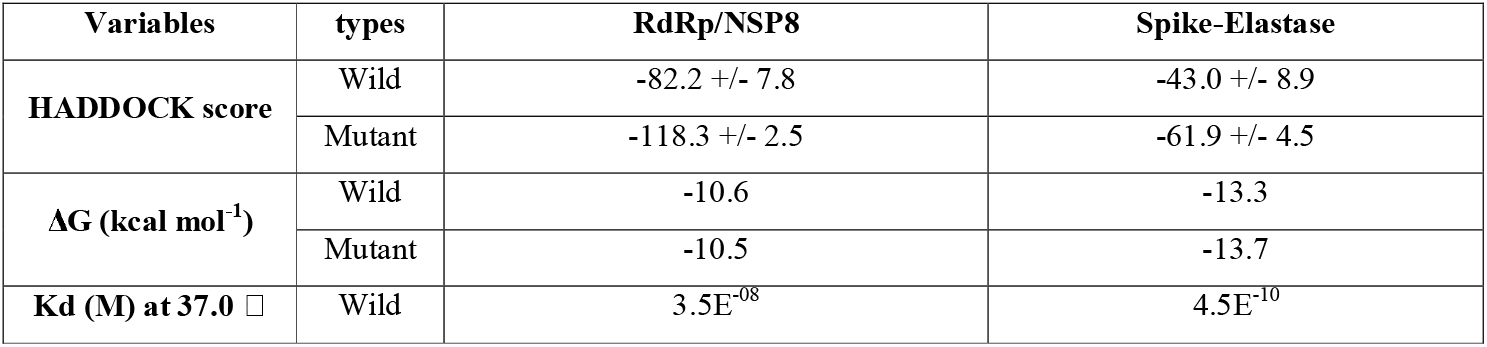
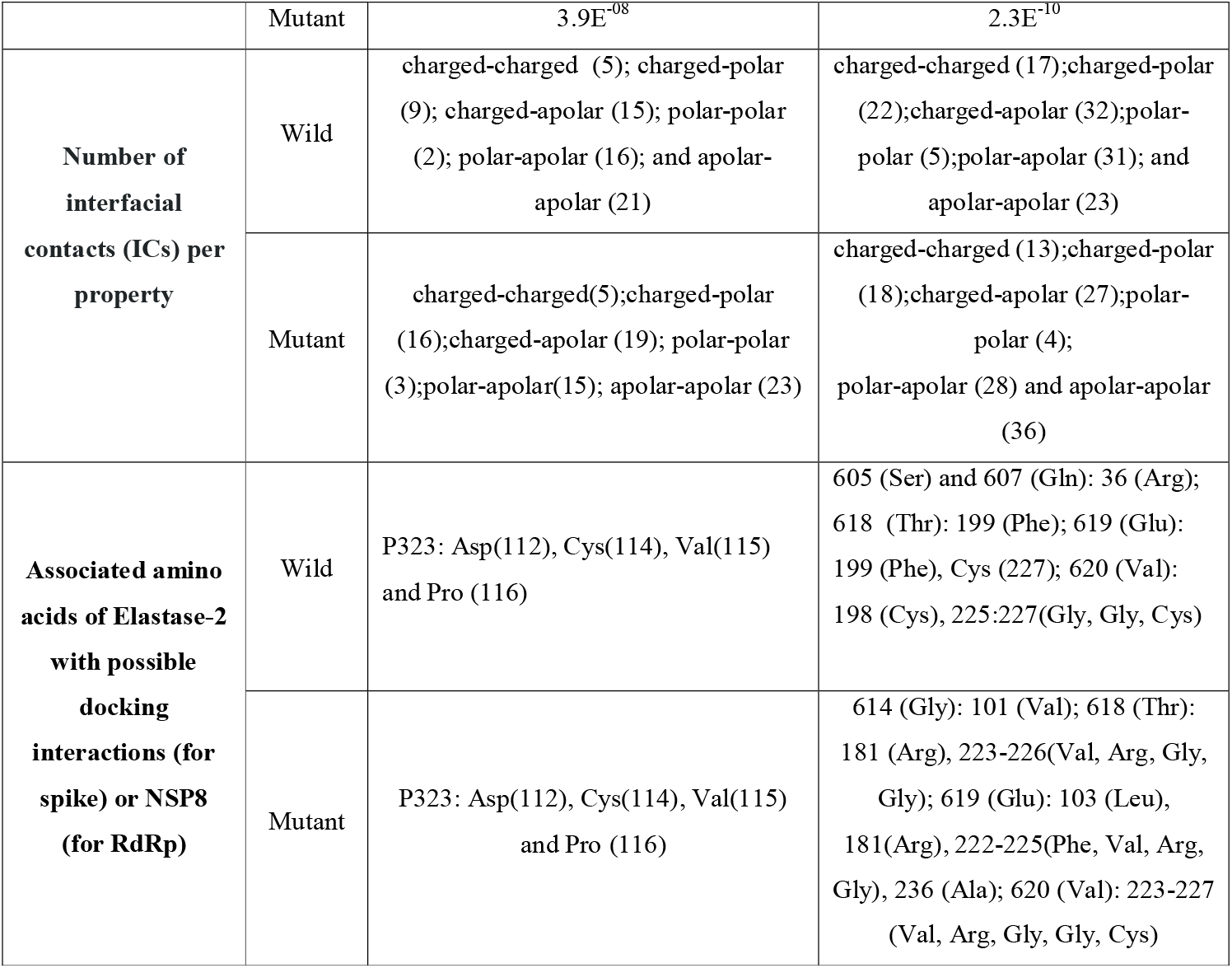
The scores of HADDOCK, PRODIGY (ΔG and Kd (M) at 37.0 □) for RdRp/NSP8 and Spike-Elastase docked complex.

**Table 2.**
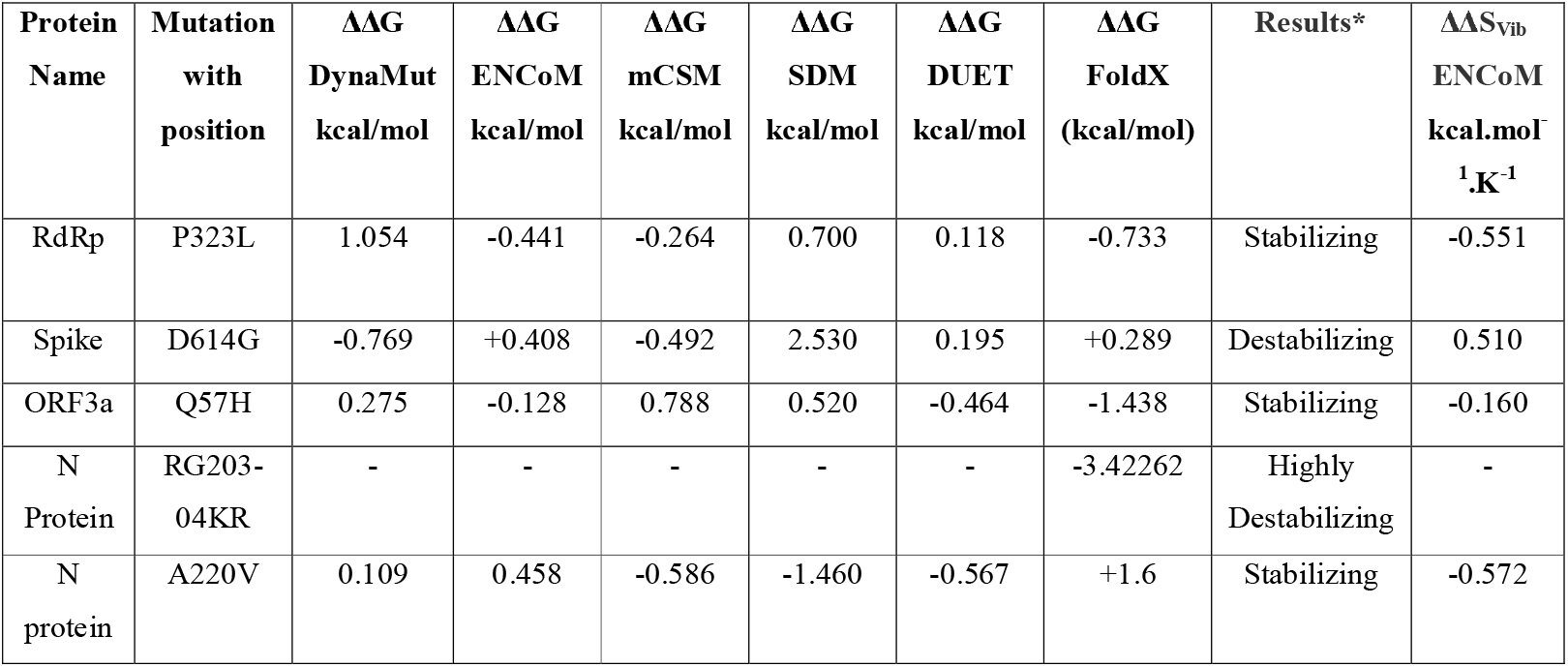
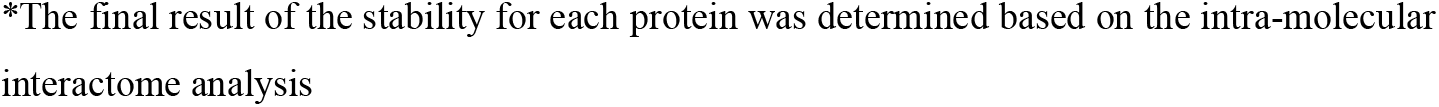
Assess the effect of mutations on structural dynamics of NSP-12/ RDRP, Spike, NS3 and N Protein of SARS CoV-2 using DynaMut. The value of ΔΔG <0 indicates that the mutation causes destabilization and ΔΔG > 0 represents protein stabilization. For ΔΔSVibENCoM, positive and negative value denotes the increase and decrease of molecular flexibility, respectively.

**Figure 3.**
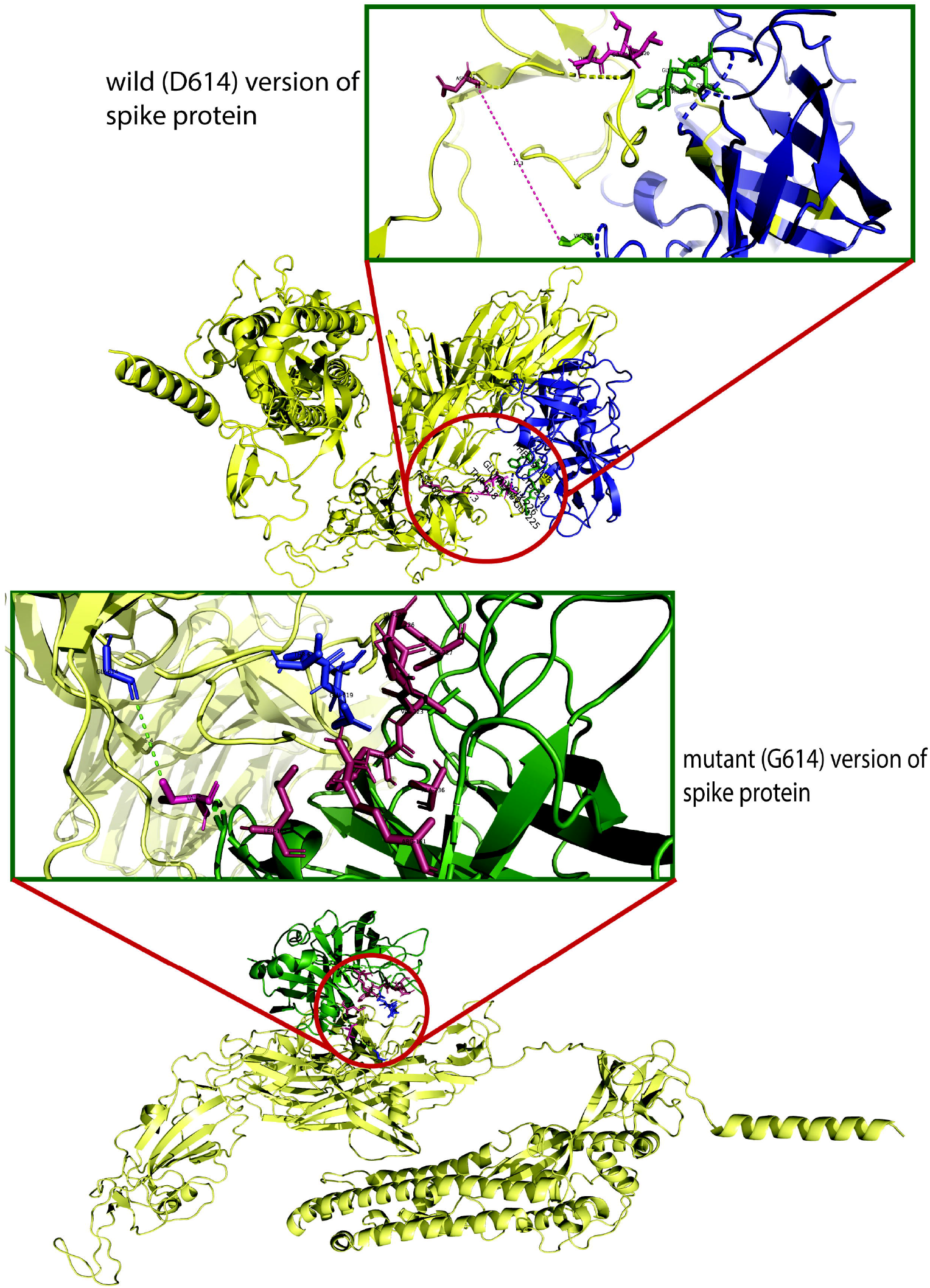
The molecular docking of wild and mutant with elastase-2. Both the (upper figure) wild (D_614_) and (lower figure) mutant (G_614_) version of S protein was shown in golden color whereas the elastase-2 docked to D_614_ and G_614_ in blue and green color, respectively. The enlarged views of the docked site were shown in separate boxes. **(a)** The possible docked residues (stick model) on the wild S protein (warm pink) and elastase-2 (green) are 618(Thr)-619(Glu)-620(Val) and 198(Cys)-199(Phe)-225:227 (Gly, Gly, Cys), respectively. The aspartic acid at 614 is 17.3°A far away from the valine (101), apparently the nearest amino acid of elastase-2 to the cleavage site (615-616). **(b)** The possible interacting residues (stick model) on the mutant S protein (blue) and elastase-2 (warm pink) are 614(Gly)-618(Thr)-619(Glu)-620(Val) and 101(Val)-103(Leu)-181(Arg)-222:227(Phe, Val, Arg, Gly, Gly, Cys), and 236 (Ala) respectively. In this case, the glycine at 614 is only 5.4°A far away from the valine (101), the nearest amino acid of elastase-2 to the cleavage site (615-616).

**Figure 4.**
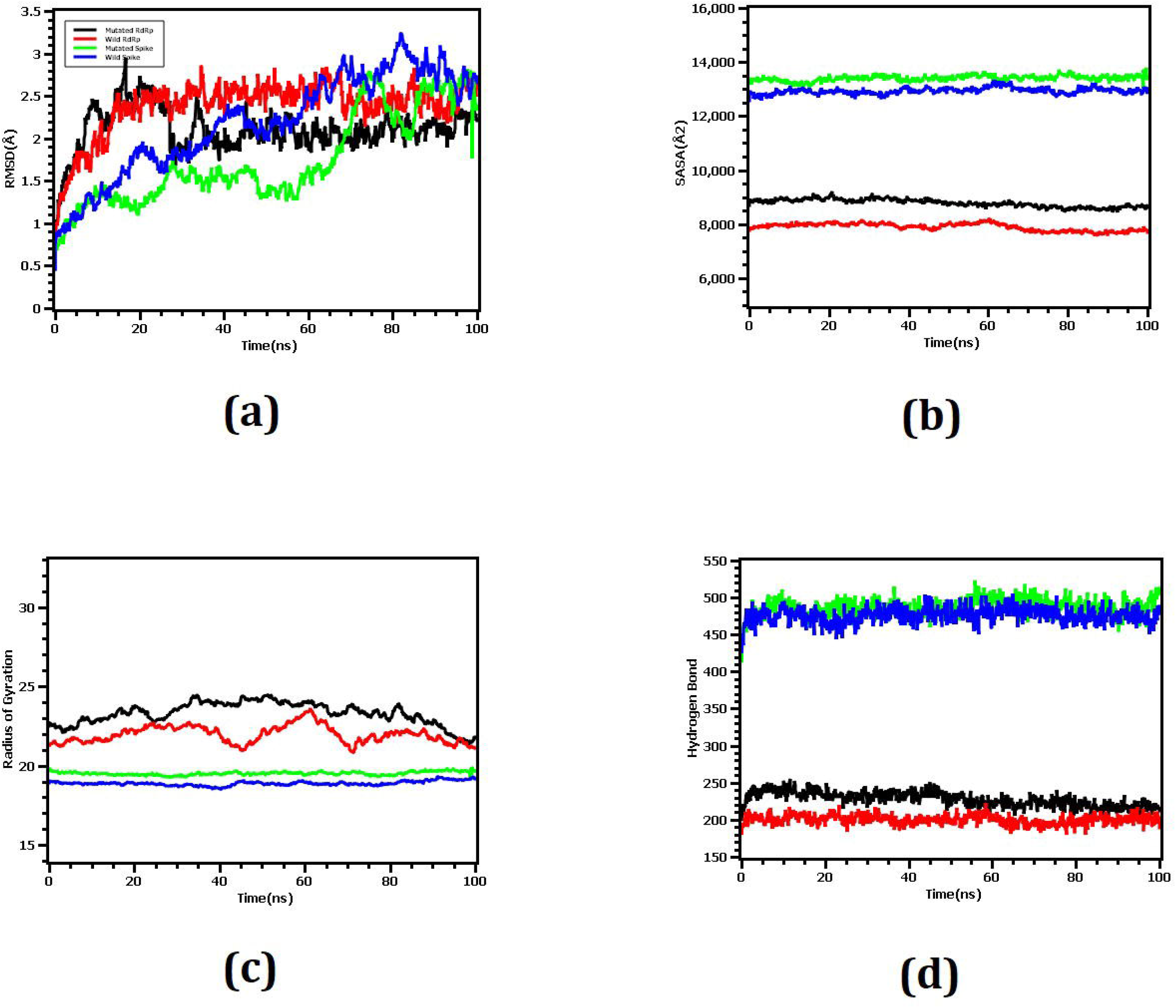
(a) Both the wild and mutated spike protein had lower RMSD profile till 60ns, then it rised and maintained steady state. Although the spike protein had higher degree of deviation in RMSD profile than RdRp but they did not exceed 3.0Å. The RMSD from demonstrated that mutant and wild RdRp protein complex has initial rise of RMSD profile due to flexibility. Therefore, both RdRp complexes stabilized after 30ns and maintained steady peak. The wild type RdRp complex had little bit higher RMSD peak than mutant RdRp which indicates the more flexible nature of the wild type. (b) The spike protein complex had similar SASA profile and did not change its surface volume and maintained similar trend during the whole simulation time. The higher deviation of SASA indicates that mutant and wild type RdRp had straight line but mutant structure had higher SASA profile which indicates the protein complex had enlarged its surface area. Therefore, mutation in RdRp protein leads to more expansion of the surface area than wild types as their average SASA value had significant difference. (c) Mutated spike exhibits little more Rg profile than the wild type which correlates with the comparative labile nature of the mutant. The higher level of Rg value defines the loose packaging system and mobile nature of the protein systems. The mutant RdRp had lower level of fluctuations and maintains its integrity in whole simulation time. The wild type RdRp complexes had higher deviations and more mobility than the mutant complex. (d) Any aberration in hydrogen bond number can lead to a higher flexibility. Therefore, the mutant and wild spike protein exhibit same flexibility in terms of H-bonding. The mutant RdRp protein had more hydrogen bonding than the wild types, but they did not differentiate too much and relatively straight line was observed for the protein.

This G^614^ amino acid replacement may have a destabilization effect on the overall protein structure (Table 2 and Figure 2a-b). The deformed flexible region at or near G^614^ is the proof of that destabilizing change (Figure 2f and supplementary figure S3). Zhang et al.^59^ explained less S1 shedding through more stable hydrogen bonding between Q^613^ (glutamine) and T^859^ (threonine) of protomer due to greater backbone flexibility provided by G^614^. Moreover, G614 mutation may increase S protein stability and participate in N-linked glycosylation at N616^36^. On the other hand, increasing the number of RBD up conformation or increasing the chance of 1-RBD-up conformation due to breakage of both intra-and inter-protomer interactions of the spike trimer and symmetric conformation will give a better chance to bind with ACE2 receptor and can also increase antibody-mediated neutralization^60^. The S1 will release from S2 more effectively in G_614_ protein by introducing glycine that will break hydrogen bond present in between the D^614^ (wild) and T^859^ (threonine) of the neighboring protomer^60,61^. Our analyses have provided the *in silico* proof of this later fact by showing that the mutated protein was more flexible than the wild type protein by missing a hydrophobic interaction between G^614^ and Phe^592^ (Figure 2g-h). This new adjunct result accorded well with Weismann et al. (2020) that G^614^ will increase overall flexibility of the mutated protein. The overall structural change may assist the mutated S protein by providing elastase-2 a better binding space and attachment opportunity onto the cleavage site (Figure 3a-b), thus providing a more stable interaction that increases the credibility of an efficient infection (Figure 1).

### 3.2 Increased Flexibility of RdRp-NSP8 Complex: Compromise Proof-Reading Efficiency with Replication Speed

The binding free energy (ΔG) of the RdRp-NSP8 complexes have been predicted to be -10.6 and -10.5 Kcalmol^-1^, respectively, in wild (P_323_) and mutated (L_323_) type that suggests a more flexible interaction for the mutated protein (Table 2). The increased number of contacts found in the L_323_-NSP8 complex (Table 1) were possibly due to slightly more hydrogen bonds, which had no considerable impact on protein flexibility (Figure 4d). Our analyses identified that proline (P^323^) or leucine (L^323^) of RdRp can interact with the aspartic acid (D^112^), cysteine (C^114^), valine (V^115^), and proline (P^116^) of NSP8 (Table 1 and Figure 5). RdRp binds with NSP8 in its interface domain (from residues alanine:A^250^ to arginine:R^365^), forming positively charged or comparatively neutral ‘sliding poles’ for RNA exit, and enhance the replication speed probably by extending the RNA-binding surface in that domain area^62,63^. Molecular dynamics of the mutated RdRp-NSP8 complex supported this by showing a more expanded surface area in the interacting site (Figure 4b) and maintained integrity throughout the simulation (Figure 4c). Besides, we did not find any interaction of NSP8 with the zinc-binding residues (H^295^, C^301^, C^306^ and C^310^) of the RdRp protein (Table 3 and Figure 5)^64,65^. Therefore, the P323L mutation within this conserved site of the RdRp interface domain may only affect the RdRp-NSP8 interaction without changing metal binding affinity.

**Figure 5.**
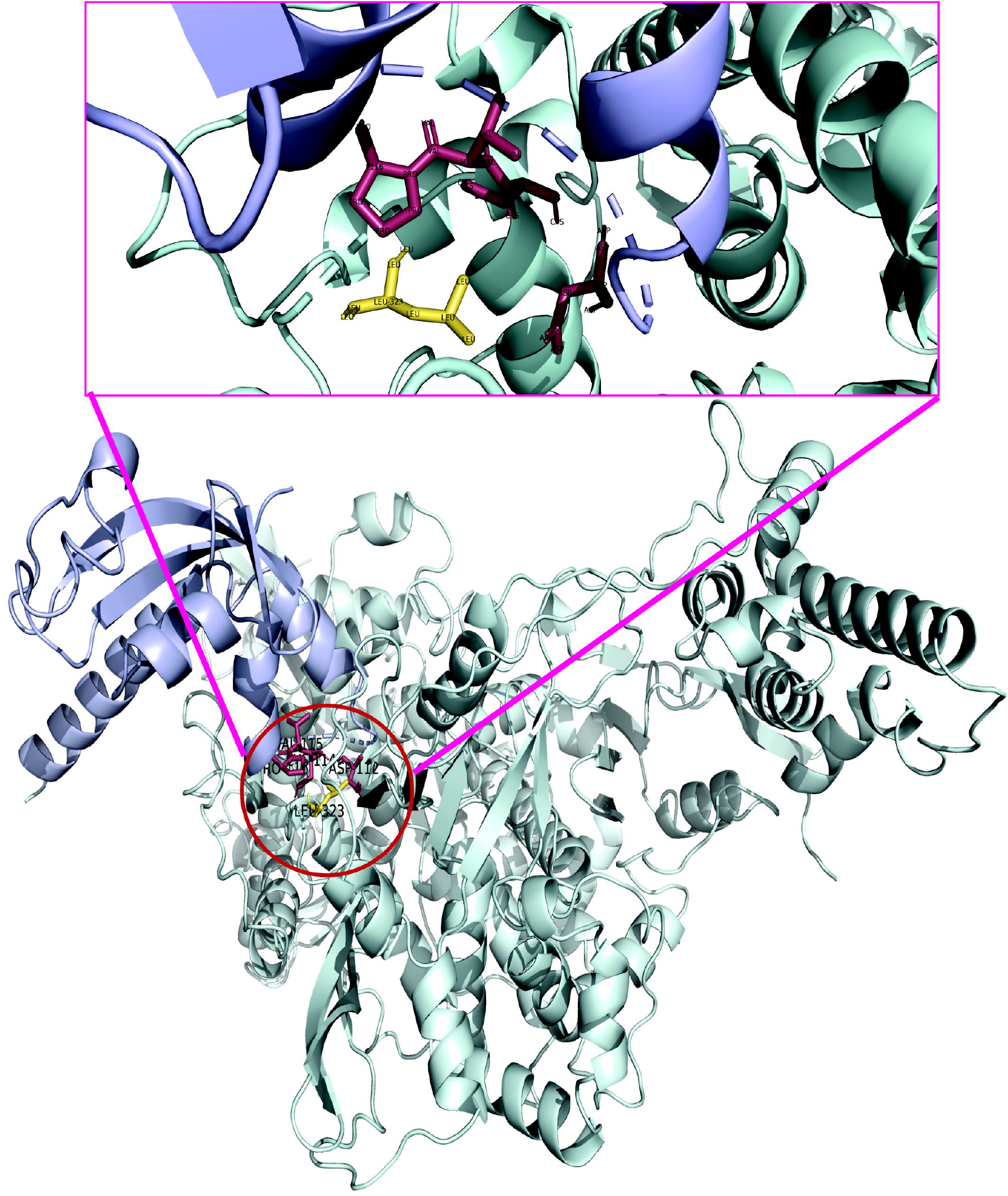
The molecular interaction of mutant RdRp with NSP8. The mutant (L_323_) RdRp (pale green) and NSP8 (light blue) are interacting as shown in center of the lower figure and an enlarged view of the docked site is presented above within a box. The leucine at 323 interacted with the Asp (112), Cys (114), Val (115), and Pro (116). The wild (P_323_) RdRp has identical docking interactions with NSP8 (table 4), thus is not presented as separate figure here.

The results from six state-of-the-art tools of protein stability suggested that mutated (L_323_) protein cannot be concluded as ‘stable’, only because of ambiguous ΔΔG estimates (Table 2), rather the interaction with the adjacent amino acid mainly defined the stability^66^. The superimposed 3D structures and secondary structure analyses showed that there was no deviation in loop/turn structure of mutated protein, even though hydrophobic leucine (L^323^) was embedded (supplementary Figure S1 and S4). To some extent, the mutation stabilized the L_323_ structure making the protein more rigid and binds less strongly with the NSP8 by expanding the interacting region. These variations may together increase the replication speed by helping exit the processed RNA genome from the RdRp groove structure more swiftly (Figure 1). The increasing replication speed might be due to the perturbation of interaction between RdRp and NSP8^62,66^, or less possibly, the complex tripartite interactions (RdRp, NSP8, and NSP14) responsible for the speculated decrease of proof-reading efficiency^4^. Thus, RdRp mutants might increase the mutation rate by a trade-off between high replication speed and low fidelity of the mutated polymerase^67^. Another possibility could be the lower proof-reading efficiency of NSP14 that was however not linked to the replication speed^4^. Analysis of our study sequences revealed that the frequency of mutation (median=8) in L_323_ mutants (n=27,364) is significantly higher (p<0.0001) than the frequency (median=6) of wild-type (P_323_) strains (n=9,815). This increased mutation rate may play a vital role in genetic drifts and provide next generations a better adaptation to adverse environments.

### 3.3 Q57H Substitution in ORF3a Viroporin: The Roles of Decreased Ion Permeability

This study has found that the replacement of glutamine (Q^57^) with positively charged histidine (H^57^) at 57 position of ORF3a transmembrane region 1 (TM1) does not change secondary transmembrane helical configuration (supplementary Figure S1), and aligned 3D structures have also shown no variation of TM1 in the monomeric state (Figure 6a). The mutant (H_57_) protein has a non-significant increase in structural stabilization and a minimal decrease in molecular flexibility (Table 2 and supplementary Figure S5). This is because of the weak ionic interaction of H^57^-C_α_ with the sulfur atom of cysteine (C^81^) that is present in TM2 and the hydrogen bond of terminal N_ζ_ of lysine (K^61^) with one of the endocyclic nitrogens of H^57^ (Figure 6b). The Q^57^ in wild-type protein forms the major hydrophilic constriction within the ORF3a channel pore^68^. Thus, further favorable increasing constrictions within the H_57_ protein channel pore due to diagonal H^57^ (TM1)-C^81^ (TM2) ionic interaction (Figure 6b) and the replacement of charge-neutral glutamine with a positively charged histidine in the selectivity filter may reduce the passing of positive ions, such as Ca^2+^, Na^+^, and K^+^, by either electrostatic repulsion or blocking^69-72^. This speculation for ORF3a mutated protein was supported by another study showing the reduction of ion permeability of Na^2+^ and Ca^2+^ through the H_57_, however, that decrease was not found statistically significant (p>0.05)^68^.

**Figure 6.**
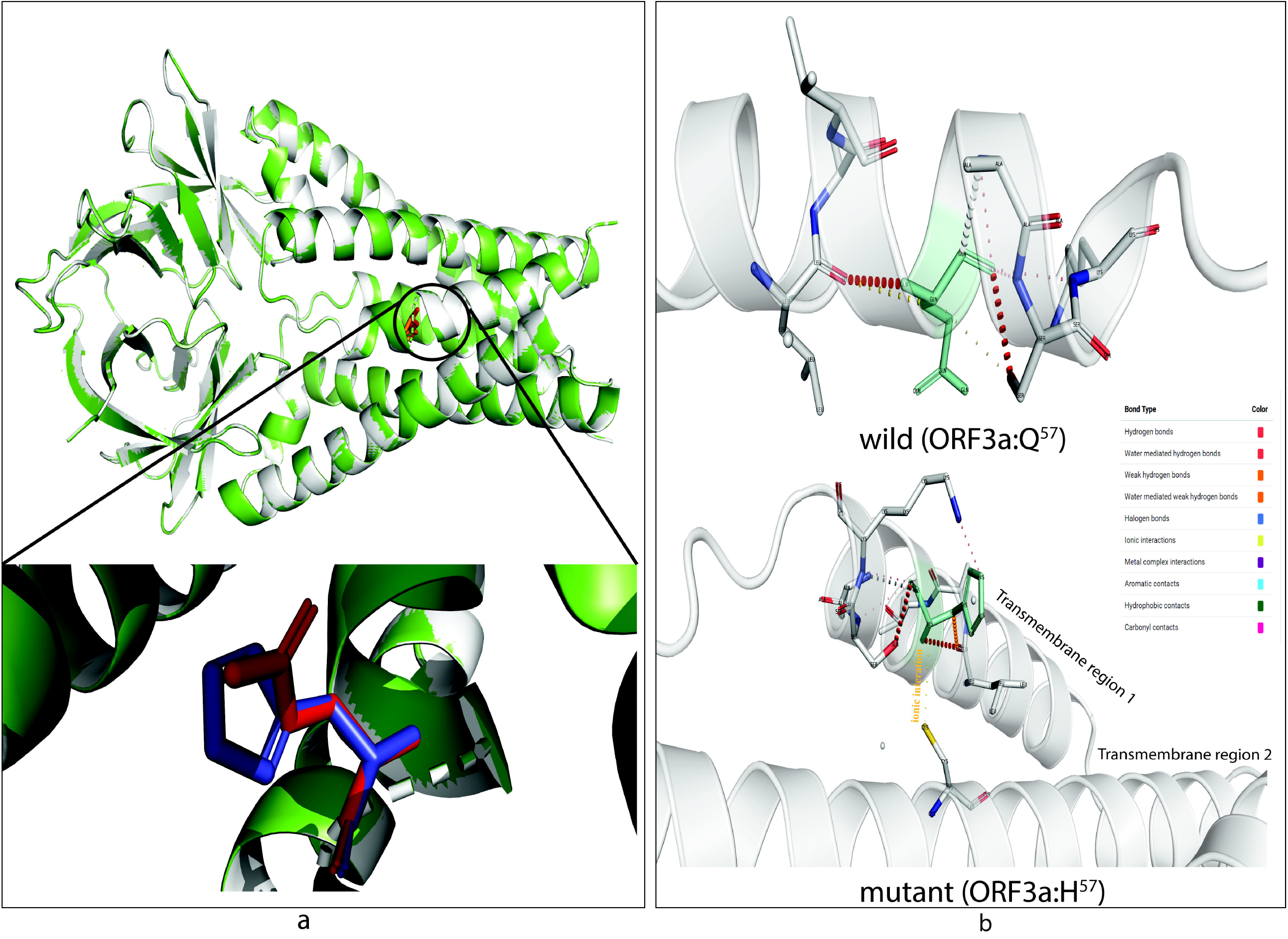
The effect on transmembrane channel pore of ORF3a viroporin due to p.Q57H mutation. **(a)** The wild (Q_57_) and mutant (H_57_) ORF3a protein are presented in light gray and green color, respectively. The structural superposition displays no overall conformation change, however the histidine at 57 position of mutant ORF3a (deep blue) has slightly rotated from glutamine at same position of the wild protein (bright red). This change in rotamer state at 57 residue may influence **(b)** the overall stability of H_57_ (upper part) over Q_57_ (lower part) because of ionic interaction of histidine (green; stick model) of transmembrane domain 1 (TM1) with cysteine at 81 (yellow stick) of TM2. The color code defined different bond types is shown in inlet.

The decrease intracellular concentration of cytoplasmic Ca^2+^ ions potentially reduces caspase-dependent apoptosis of the host cell^73^, mainly supporting viral spread without affecting replication^21^ as shown in Figure 1. Moreover, the ORF3a can drive necrotic cell death^74^ wherein the permeated ions into cytoplasm^75^ and the insertion of ORF3a as viroporin into lysosome^76^ play vital roles. The H_57_ mutant may thus decrease pathogenicity and symptoms during the early stages of the infection, i.e., reducing ‘cytokine storm’ in the host^77^. Besides, ORF3a was proved to affect inflammasome activation, virus release, and cell death, as shown by Castaño-Rodriguez et al. (2018)^78^ that the deletion of ORF3a reduced viral load and morbidity in animal models.

Even though similar proteins of ORF3a have been identified in the sarbecovirus lineage infecting bats, pangolins, and humans^79^, only one pangolin derived strain from 2017 in Guangxi, China contains H^57^ residue as shown by mutation analyses (supplementary Figure S5), and also reported by Kern et al. (2020)^68^. The presence of this mutation in pangolin could be an accidental case or might explain its impact on modulating host-specific immune response, which needs functional experimental verification. A possible explanation behind that presence could be the more adaptive nature of the virus towards reverse transmission by being less virulent, i.e., from human to other animals, as observed in recent reports^80,81^.

### 3.4 N Protein Mutation: Augmenting Nucleocapsid Stability and Exerting Miscellaneous Effects

Our study has observed that the combined mutation (N: p.RG203-204KR) causes no conformational change in secondary and 3D structures (Figure S1 and Figure 7, respectively) of the conserved SR-rich site (184 → 204) in the linker region (LKR: 183 → 254) of the N protein (supplementary Figure S7), but there is a minor alteration among buried or exposed residues (supplementary Figure S1c and Figure 8b). The superimposed 3D structures showed structural deviation, rather at ^231^ESKMSGKGQQQQGQTVT^247^ of the LKR (Figure 7), corresponding to the high destabilization of the mutated (KR_203-204_) protein (Table 2).

**Figure 7.**
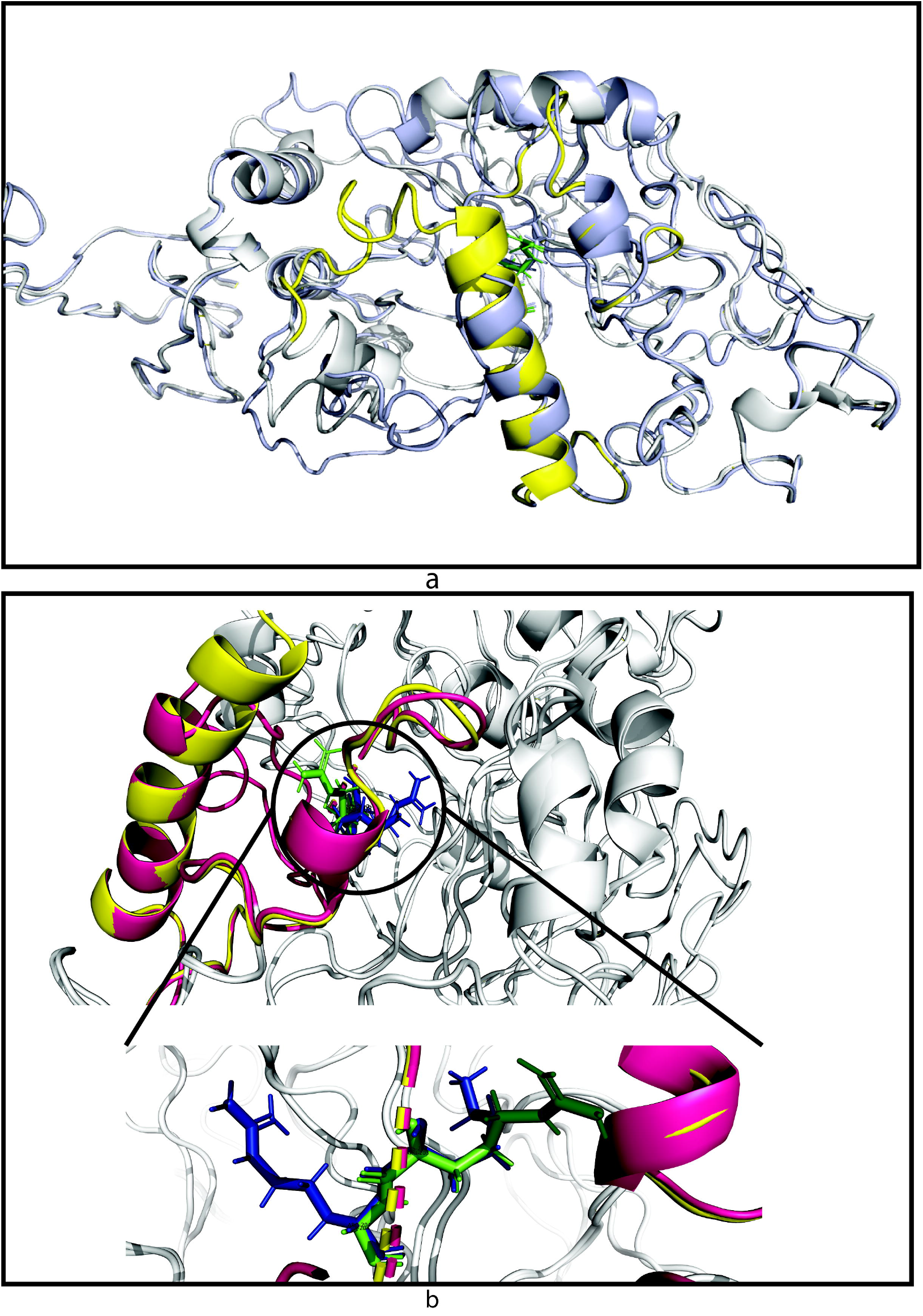
Structural superposition of wild and mutant N protein. The light grey color represents both wild (RG_203-204_) and mutant (KR_203-204_) N protein. The linker region (LKR: 183-247 amino acids) of wild (RG_203-204_) and mutant (KR_203-204_) are in pale yellow and warm pink color, respectively. **(a)** The aligned structures showed a highly destabilizing (Table 3) conformational change from 231 to 247 amino acids within LKR. Other regions of the N protein, especially the SR-rich region (184-204 amino acids) where the mutations occur, do not change. **(b)** A more emphasized look into the SR-rich and mutated sites (RG203-204KR) of wild and mutant N protein represent slight deviation in the Ser (197) and Thr (198) while only glycine (green) to arginine (blue) substitution at 204 position shows changing at rotamer state. The enlarged view is shown in the bottom part.

Impedance to form particular SR-motif due to RG→KR mutation might disrupt the phosphorylation catalyzed by host glycogen synthase kinase-3^82^. After virus enters into the cell, this synchronized hypo-phosphorylation of KR_203-204_ protein should make the viral ribonucleoprotein (RNP) unwind in a slower but more organized fashion that might have an impact upon translation and immune-modulation^83-85^. In KR_203-204_, replacement of glycine with arginine may increase the nucleocapsid (N protein-RNA complex) stability by forming stronger electrostatic and ionic interactions due to increased positive charge^86,87^. Besides, the more disordered orientation of the downstream LKR site^83^ and highly destabilizing property of KR_203-204_ may assist in the packaging of a stable RNP^88,89^. N protein also utilizes the dynamic nature of the intrinsically disordered linker region (LKR) that controls its affinity towards M protein, self-monomer, 5’UTR, and cellular proteins^90-92^. The phosphorylation at the LKR site may play an essential role to regulate these interactions^86^. These plausible interactions and impact upon mutations are depicted in Figure 1.

### 3.5 Silent Mutations may not be Silent

The C241T of 5’UTR (untranslated region), a single nucleotide ‘silent’ mutation, is located at the UUCGU pentaloop part of the stem-loop region (SLR5B). This pentaloop of 5’UTR remains unchanged and maintains a particular structure with a potential role in viral packaging^91,93^. The RNA secondary structural analysis in our study predicted that there is no change in the 241T structure (supplementary Figure S8a). However, C241T is present just upstream to the ORF1a start codon (266-268 position) and may be involved in differential RNA binding affinity to the ribosome and translational factors^94^.

In the case of multi-domain NSP3 (papain-like protease), we have observed superior stability of the RNA after gaining the synonymous mutation 3037C<T (C318T) where wild and mutant RNA structure has -151.63 and -153.03 Kcal/mol, respectively (Supplementary Figure S7b-c). A more stable secondary structure of (+)-ssRNA as observed in the mutated NSP3 protein coding sequence corresponds to the slower translational elongation that generally contributes to a range of abnormalities resulting in low translation efficiency affecting posttranslational modifications as a part of protein regulation^95^. This silent mutation is located within the flexible loop of the NSP3 ubiquitin-like domain 1 (Ubl1). In SARS-CoV, Ubl1 was reported to bind with single-stranded RNA containing AUA patterns and interact with the nucleocapsid (N) protein^96,97^. Besides, Ubl1 was likely to bind with several signature repeats in 5′-UTR in SARS-CoV-2 genome^98^. Finally, change in T445C in leader protein may not cause any change in expression or others since the structure (data not shown) and energy are same -172.34 kcal/mol. Figure 1 represents the overall possible scenario due to these silent mutations.

### 3.6 Epistatic Effects of the Co-occurring Mutations on Viral Replication and Transmission: A Plausible Hypothesis

The co-occurring mutations, as defined by the presence of simultaneous multi-site variations in the same or different proteins or in the genome, have provided new insights into the dynamic epistatic network by employing differential molecular interactions. The epistatic effects of the mutations were reported to control viral fitness and virulence through modulating replication cycle and virus-host interactions, as observed before for Influenza and Ebola virus^99-104^. For instance, the detrimental effect of R384G on influenza A fitness was overcome by the co-occurring mutation E375G^100,103^, and co-occurring mutations at the antigenic sites of influenza hemagglutinin can also drive viral evolution^102,104^. The correlation of the co-occurring GP-L mutations affect Ebola virus virulence and thus case fatality rate^101^. The co-occurring mutations of the major SARS-CoV-2 clades discussed in this work showed epistatic link^22,23^, and positive selection pressure except for the synonymous mutations^22,24,25^ that were also shown in Observable site (https://observablehq.com/). We propose here a hypothesis (model) on how the co-occurring mutations of SARS-CoV-2 might influence replication and transmission fitness of the major clade strains.

Between two important G clade-featured co-occurring mutations, the p:D614G of the S protein and p:P323L of RdRp, we speculated no interlinked functional relationship (Figure 1) and the sequence based prediction showed no potentially significant epistatic link as well^23^. These seemingly unrelated mutations can cumulatively escalate the infectiousness of the virus as a result of higher viral load and shorter burst time. The S:p.D614G might assist in rapid entry into the host cells with an efficient elastase-2 activity and higher transmissibility and/or aggressiveness of G_614_ mutant^19,20^ and could be related to elastase-2 or human neutrophil elastase (hNE) concentration during inflammation^105^. Besides, higher levels of functional S protein observed in G614 strains can increase the chance of host-to-host transmission^59^. The RdRp:p:P323L may rather boost up the replication by a faster RNA processing (exiting) that can open up the avenue to generate strains with significantly (p<0.0001) higher number of mutations. This increased mutation rate in L_323_ mutants can surpass the constant proof-reading fidelity^106^ and evolve into a greater number of variants within a population (Figure 1), that might help adapt more quickly in adverse climatic condition, evade the immune response, and survive within different selective pressure^107,108^.

NSP3 is a scaffolding protein for the replication-transcription complex, and the possible change in its structure may affect the overall dynamics of viral replication^96,97^. P323L mutation of RdRp may change binding affinity to the Ubl1 region of NSP3^109^ (Figure 1). Significant epistatic links of NSP3:C3037T with spike and RdRp mutations was also reported by Zeng et al. (2020)^23^. In contrast, we could not predict any possible association of the 5’
sUTR:C241T mutation with the S, RdRp, and NSP3 mutated proteins as also shown by sequence analysis in Zeng et al. (2020)^23^. The rapid within-host replication and modified replication dynamics might be correlated with the fitness of G clade strains^110^.

The mutant N protein may have an impact on viral replication and transcription, like other coronaviruses^92^, through the binding with NSP3 protein that is linked to RdRp centered replication complex. The N protein can also affect the membrane stability by yet uncharacterized interaction with the M protein, which could ultimately produce more stable virion particles^111-113^. A stronger N protein-RNA complex provokes slower intracellular immune response^84^, and at the same time, can still remain highly contagious and aggressive because of the concurrent presence of G clade-featured S protein and RdRp mutations (Figure 1). The GR strains could hence attain a plausible advantage over G and GH by a more orchestrated, delicately balanced synergistic effects on replication and transmission fitness. These epistatic effects might have increased the fitness by hiding the virus from cellular immunity of the host and increasing stability in the environment, i.e., more transmissible through air and surface. Conversely, we have not found any literature for even other coronaviruses that correlated the ORF3a: p.Q57H with rest of the co-occurring mutations. The H_57_ mutant, possibly linked to the mild or asymptomatic cases, may allow the silent transmission and increase the chance of viral spread by lowering the activation of inflammatory response (Figure 1), such as reduced viral particle release and cytokine storm^21,114,115^.

The GV strains featuring an A222V mutation in the S protein along with other mutations of the clade G was reported to have probably no effect on the viral transmission, severity, and escaping antibody due to its structural position, rather superspreading founder events after lifting up of travel restriction in Europe and lack of effective containment might be the cause of its spreading within Europe^17,116^. There was, however, speculation about the effect of this mutation on immunogenicity since computational analysis predicted its location within T-cell target region^117,118^. The A220V mutation in the N protein of the GV clade showed a slightly more stable formation of the mutated N protein linker region (Table 2) with no change in the chemical properties, that might affect RNA binding affinity^88^, but how this change might give an advantage for the GV strains is a question for further experiments. However, different mutations at positions 220 in N found in other major lineages showed no clear evidence of phenotypic consequence (Hodcroft et al., 2020). There was also no epistatically linked pairing between GV clade co-occurring mutations^22,23^. Altogether, the co-occurring mutations of GV strains might not affect transmission fitness.

Vaccine inequity, immunocompromised patients, and a tremendous number of hosts are now frequently introducing variants with mutations in the receptor binding domain (RBD) of spike protein, i.e., lineage B.1.1.7, a variant of concern under GR clade, consists of 17 mutations (14 nonsynonymous and 3 deletions) in spike, N, ORF1ab, and ORF8 proteins, as well as, 6 silent mutations^119,120^. The newly added mutations in this lineage on top of the original GR clade-featured ones in the genome might play most crucial role both in increasing transmission fitness and a slightly reduced neutralization to antibody by showing epistatic effects^121121,122^. Other emerging variants (B.1.617.*, B.1.351, B.1.258, etc.) with additional co-occurring mutations are also the descendants of these major clades. Future studies are necessary to investigate the roles of the ‘mutation package’ present in each of these variants of concern/interest.

## 4. CONCLUSION

In 2020, the course of COVID-19 pandemic was dominated by the G, GH, GR, and GV clades. The G clade-featured co-occurring mutations might increase the viral load, alter immune responses in host and modulate intra-host virus genome plasticity that arises the speculation of their probable role in frequent transmission. The GR and GH clade mutant with the signature mutation, respectively, in nucleocapsid and ORF3a protein might contribute to immune response of the host and viral transmission. The GV strains however could have spread quickly by superspreading events with no apparent epistatic effect. Therefore, the fitness of SARS-CoV-2 may increase in terms of replication and transmission where viral strains are always giving their spread capacity within a population the top priority by calibrating the infection cycle. However, further *in vivo* and *ex vivo* studies and more investigations are required to prove and bolster this hypothesis.

## Data Availability

All the sequence data were taken from the GISAID (https://www.gisaid.org/) and RCSB PDB (https://www.rcsb.org/) as mentioned in the methodology section. We provide all the necessary information such as accession numbers, date-based data source for helping readers and reviewers to check the authencity of the work.

## Data Availability

All the sequence and structural data were taken from the GISAID (https://www.gisaid.org/) and RCSB PDB (https://www.rcsb.org/) as mentioned in the methodology section. We provide all the necessary information such as accession numbers, date-based data source for helping readers and reviewers to check the authenticity of the work.

## Acknowledgments

We would like to acknowledge the team at GISAID for creating SARS-CoV-2 global database. The funding of the research was provided by Jashore University of Science and Technology. We appreciate to the Microbial Genetics and Bioinformatics Laboratory of University of Dhaka for the support of the high-performance computer access. We thanked M. Shaminur Rahman for their technical assistance in protein structure stability analyses.

## Biographical Note

A. S. M. Rubayet Ul Alam (ASMRUA) is an assistant professor in the Department of Microbiology, Jashore University of Science and Technology. His research mainly focuses on molecular biology, data analysis, and bioinformatics.

Ovinu Kibria Islam (OKI) is an assistant professor in the Department of Microbiology, Jashore University of Science and Technology. His research activity is focused on molecular biology and big data analysis.

Md. Shazid Hasan (MSH) is an assistant professor in the Department of Microbiology, Jashore University of Science and Technology. His research activity is focused on molecular biology and bioinformatics.

Mir Raihanul Islam (MRI) is the Senior Research Associate of BRAC James P Grant School of Public Health, BRAC University who is expert in statistical analysis and epidemiological study and is currently working on many active researches related to public health.

Shafi Mahmud (SM) is working as a Master’s thesis student in the department of Genetic Engineering and Biotechnology, University of Rajshahi, Rajshai-6205, Bangladesh.

Hassan M. Al□Emran (HMA) is the Chairman in the Department of Biomedical Engineering, Jashore University of Science and Technology, Jashore-7408, Bangladesh. His research mainly focuses on clinical microbiology and infectious diseases.

Dr. Iqbal Kabir Jahid (IKJ) is a Professor and Chairman in the Department of Microbiology, Jashore University of Science and Technology. His research mainly focuses on molecular biology.

Keith A. Crandall (KAC) is a Professor in the Department of Biostatistics & Bioinformatics and the director of Computational Biology Institute, Director of Genomics Core, Milken Institute School of Public Health, The George Washington University, Washington, DC, USA.

M. Anwar Hossain (MAH) is the director of Genome Center, and Vice Chancellor of Jashore University of Science and Technology. He is an expert in molecular biology, virology and vaccine development.

## Authors’ Contribution

IKJ, OKI and ASMRUA hypothesized about the work. ASMRUA performed the sequence analysis part after OKI and MSH compiled the dataset. The Python coding, structural (both RNA and protein), and protein docking were done by ASMRUA. MSH predicted protein structure in I-TASSER and performed stability analysis in DynaMut. SM performed the molecular dynamics study. HMA performed the statistical analysis. HMA then reviewed and organized the manuscript expertly. IKJ, KAC and MAH supervised, suggested and revised the write-up to produce the final draft.

