## supplementary for "Dominant Clade-featured SARS-CoV-2 Co-occurring Mutations Reveals Plausible Epistasis: An *in silico* based Hypothetical Model"

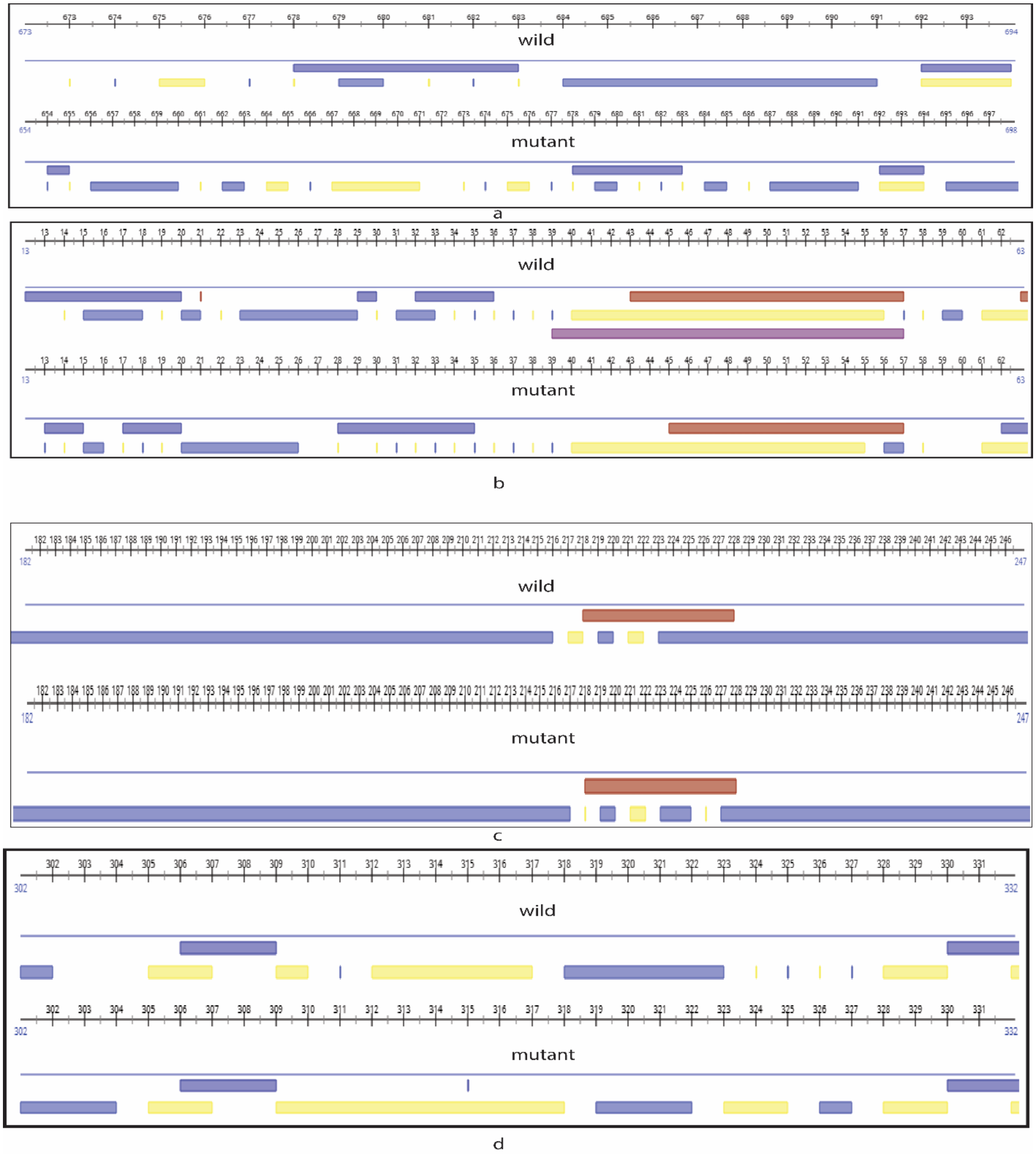


**Supplementary Figure 1. Secondary structure prediction of (a) S protein, (b) ORF3a, (c) N protein, and (d) RdRp.** The wild and mutant type was presented for each protein. Here we focused on the mutation position in the protein. The red and blue colors in the upper bar means helix and strand structures where the blue and yellow colors represent exposed and buried region, respectively. The blank space means loop and intermediate region.

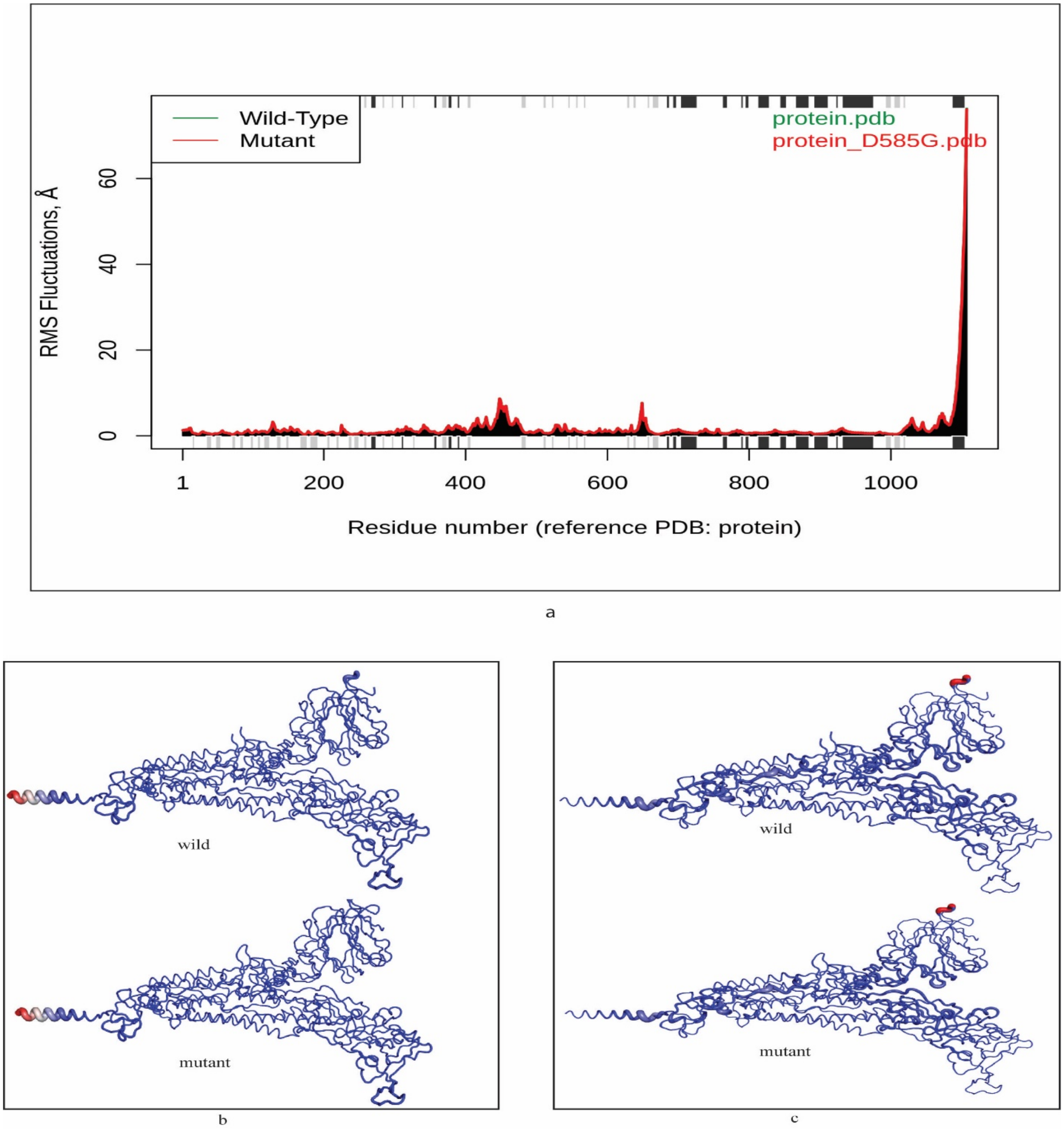


**Supplementary Figure 2. Deformation and Fluctuations comparison between wild and mutant S proteins. (a)** Ensemble normal mode analysis of wild ($D_{614}$) and mutant ($G_{614}$) types based on respective 3D structures and then aligned to compare the root-mean square (RMS) fluctuations in angstrom (°A). Helices and strands are shown in both top and bottom of the images in gray and black colors, respectively. **(b)** Atomic fluctuation and **(c)** deformation energy provide the amplitude of the absolute atomic motion and a measure for the amount of local flexibility in the proteins where the magnitude is represented by thin to thick tube colored blue (low), white (moderate) and red (high). We found no variation in the fluctuation and deformation energy between the wild ($D_{614}$) and mutant ($G_{614}$) types.

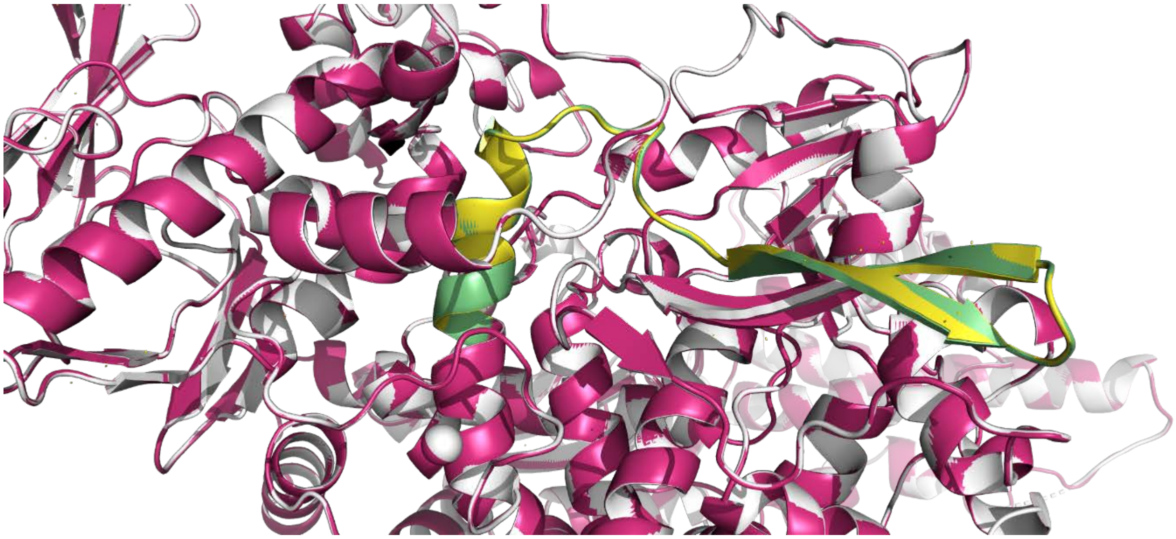

**Supplementary Figure 3. Superimposition of wild type ($P_{323}$) RdRp on mutant protein ($L_{323}$).** The overlapping of wild on the mutant RdRp showed no structural deviation. The wild and mutant RdRp were shown gray and pink colors where the specific region (310-340 amino acids) of the $P_{323}$ and $L_{323}$ were colored in yellow and green, respectively.

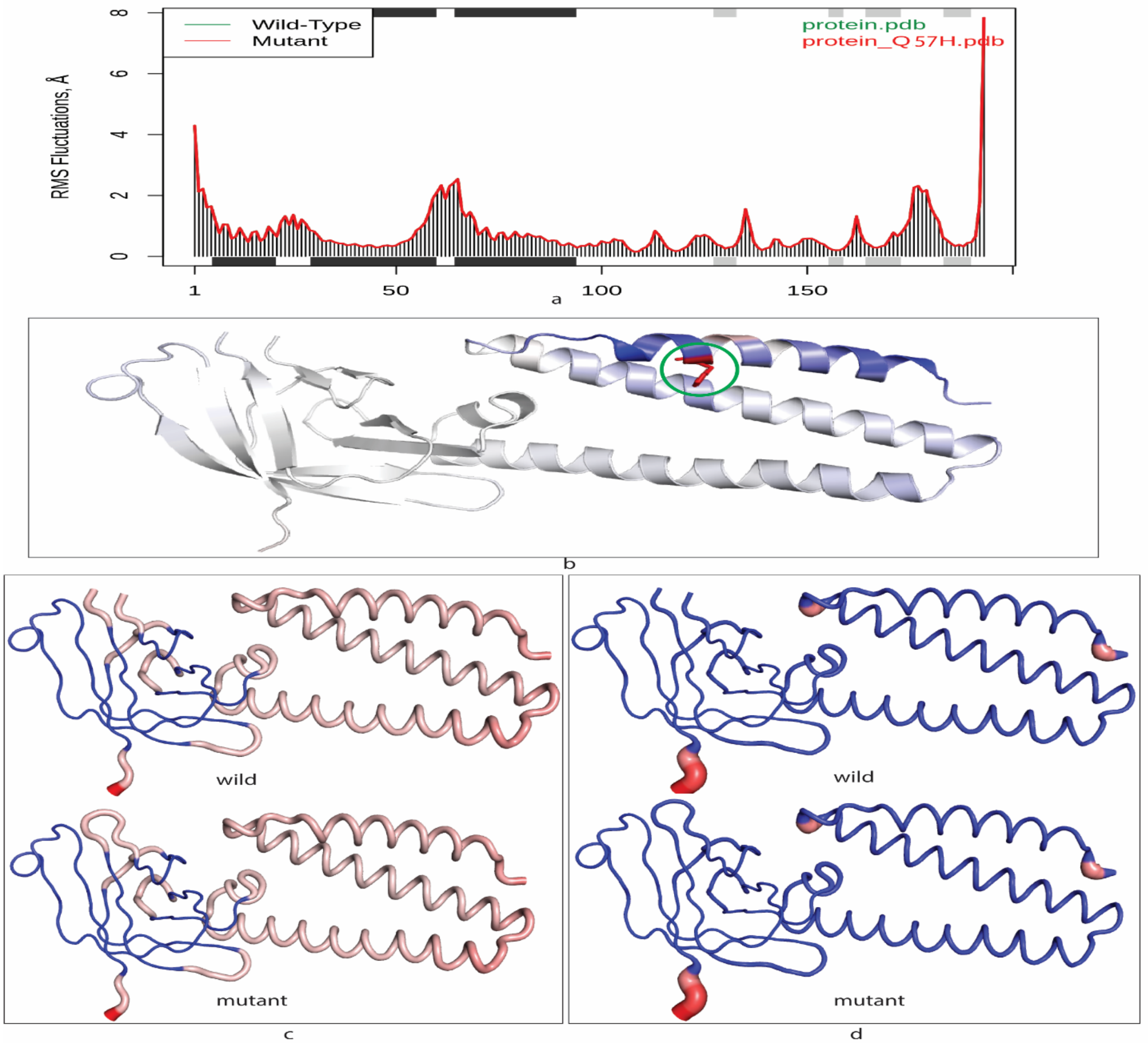


**Supplementary Figure 4. Deformation, Fluctuations, and Vibrational Entropy Energy change between wild and mutant ORF3a proteins. (a)** Ensemble normal mode analysis of wild ($Q_{57}$) and mutant ($H_{57}$) types based on respective 3D structures and then aligned to compare the root-mean square (RMS) fluctuations in angstrom (°A). Helices and strands are shown in both top and bottom of the images in gray and black colors, respectively. **(b)** The vibrational entropy change upon mutation where blue color mutation site represents a rigidification or stability of the ORF3a mutant protein structure. The amino acid $H^{57}$ and its surrounding region showed higher stability (less flexibility). **(c)** Atomic fluctuation and **(d)** deformation energy provide the amplitude of the absolute atomic motion and a measure for the amount of local flexibility in the proteins where the magnitude is represented by thin to thick tube colored blue (low), white (moderate) and red (high). We found no variation in the fluctuation and deformation energy between the wild ($Q_{57}$) and mutant ($H_{57}$).

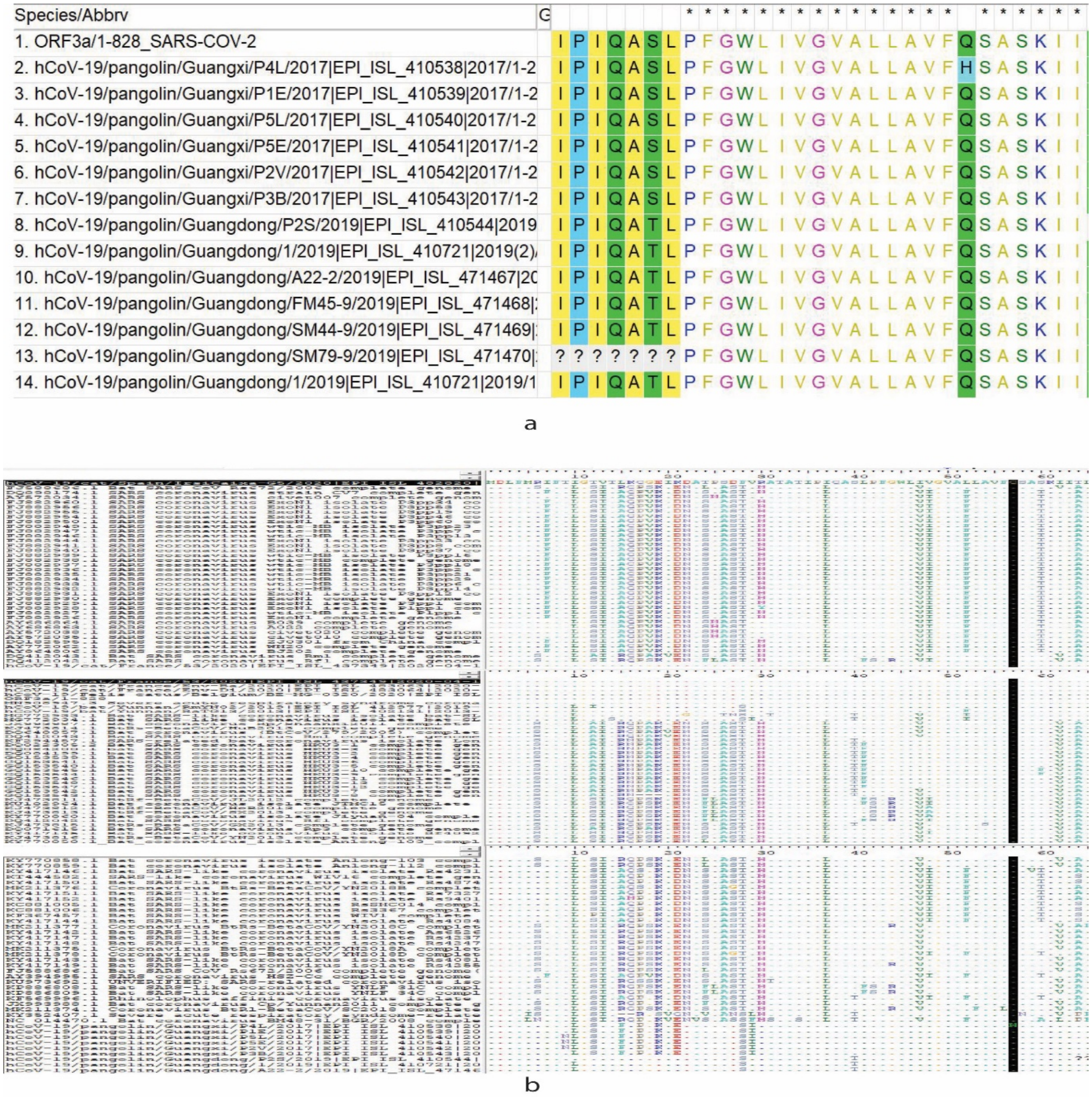


**Supplementary Figure 5. Mutation analysis showing the transmembrane region 1 (41-LPFGWLIVGVALLAVFQSASKII-63 amino acids) of SARS-CoV-2 and other related viruses of sarbecovirus lineages. (a)** The pangolin coronavirus ORF3a protein alignment with the SARS-CoV-2 showed that only P4L strain from Guangxi, China in 2017 has the $H^{57}$ mutation. Other amino acids of TM1 are not changed for pangolin viruses. **(b)** Nine out of 23 amino acids of ORF3a are conserved for the sarbecovirus lineage. These are: F(43), W(45), L(46), G(49), V(50), A(51), V(55), K(61), and I(63). The black color line along the column showed 57 position.

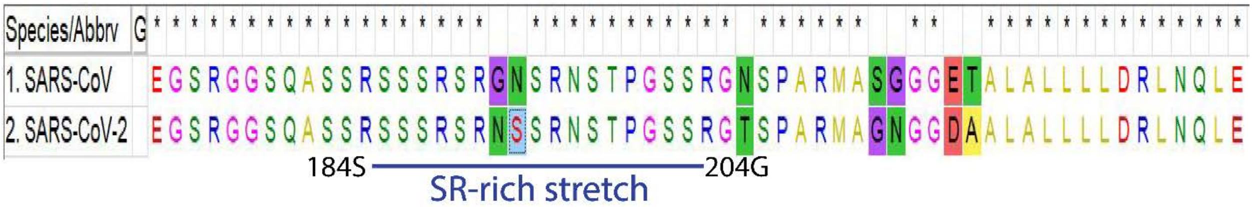


**Supplementary Figure 6. Aligned SR-rich region of SARS-CoV and SARS-CoV-2 reference sequence.** The region contained only two mutations that shows the relatedness of this region and functional report of one can be deduced from another.

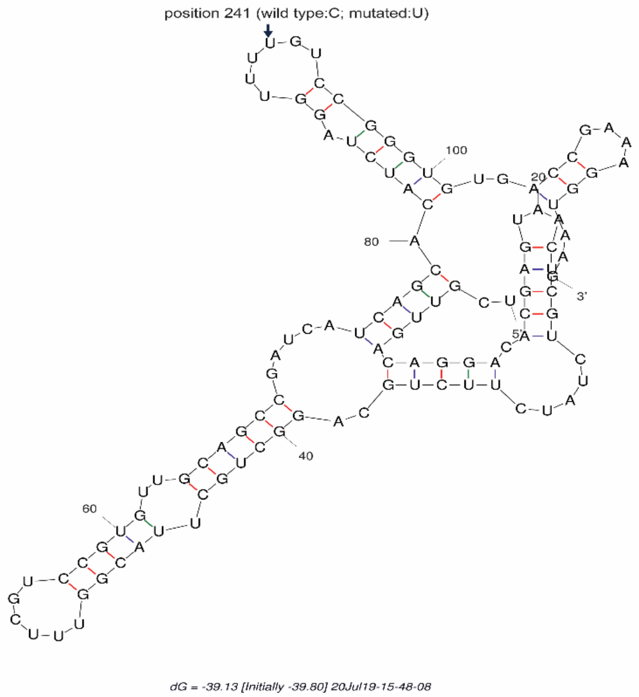


(a) 5' UTR mutant type structure

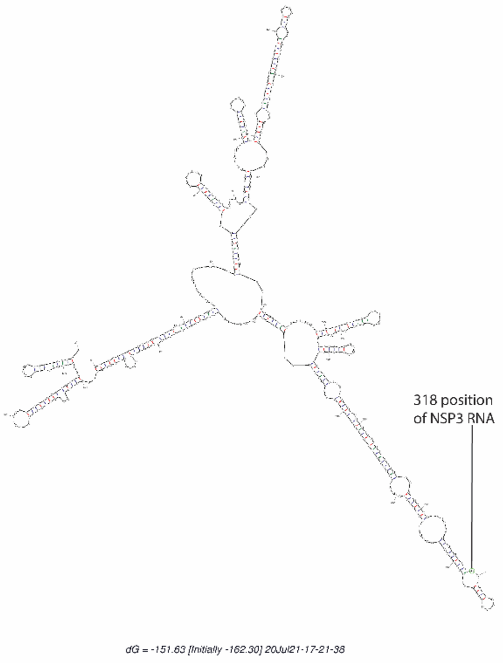


(b) Wild NSP3 RNA

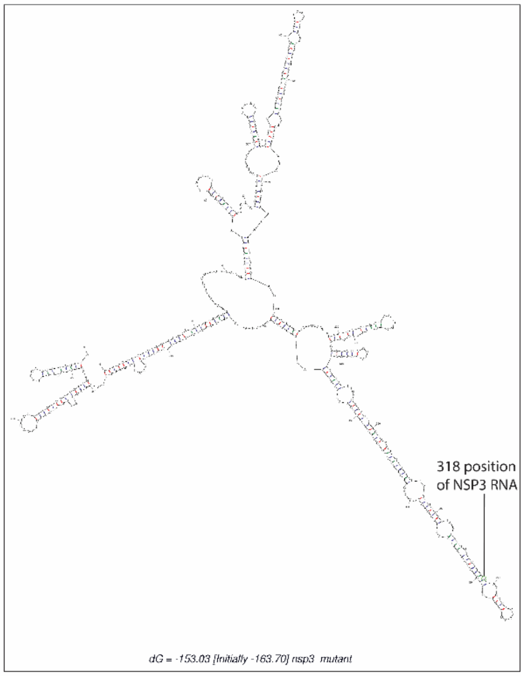


(c) Mutant NSP3 RNA

**Supplementary Figure 7. RNA secondary structures of mutant 5'-UTR (241T) and NSP3s. (a)** RNA secondary structure of 5'-UTR (151-267 positions) was determined by the Mfold web server. The changing position at 241 (91 in this image) showed 'U' instead of 'C' as found in wild-type. The change has no impact upon native secondary structure of stem-loop region 5B (SLR5B). **(b)** RNA secondary structure of NSP3 (1-500 positions) was presented in the figure. The mutated position is at 318, and there is a change from loop to stem due to this mutation (C>T) from wild type. From thermodynamic point of view, we found that the wild and mutant RNA structure have -151.63 and -153.03 Kcal/Mol stating the superior stability of the mutated type.

| Week | No. of Cas | No. of Dea | Death rate | G group (% | GH Group | GR Group | GV group (%) |
|---|---|---|---|---|---|---|---|
| Week 1 | | 0 | | 0.00% | 0.00% | 0.00% | 0.00% |
| Week 2 | | 0 | | 0.00% | 0.00% | 0.00% | 0.00% |
| Week 3 | | 0 | | 0.00% | 0.00% | 0.00% | 0.00% |
| Week 4 | | 0 | | 0.00% | 0.00% | 0.00% | 0.00% |
| Week 5 | 1732 | 0 | 12.99% | 1.45% | 0.00% | 0.00% | 0.00% |
| Week 6 | 11759 | 225 | 3.84% | 2.88% | 0.00% | 0.00% | 0.00% |
| Week 7 | 20167 | 451 | 3.76% | 4.73% | 1.78% | 0.00% | 0.00% |
| Week 8 | 11303 | 759 | 6.08% | 0.00% | 4.13% | 0.83% | 0.00% |
| Week 9 | 7382 | 687 | 4.86% | 8.26% | 3.21% | 0.46% | 0.00% |
| Week 10 | 7806 | 359 | 6.93% | 22.01% | 3.75% | 16.89% | 0.00% |
| Week 11 | 16638 | 541 | 11.58% | 23.89% | 13.80% | 23.02% | 0.00% |
| Week 12 | 43940 | 1926 | 14.06% | 19.69% | 31.85% | 13.49% | 0.00% |
| Week 13 | 124627 | 6177 | 12.34% | 20.78% | 34.91% | 15.65% | 0.04% |
| Week 14 | 301883 | 15382 | 9.83% | 22.58% | 30.87% | 21.43% | 0.00% |
| Week 15 | 440476 | 29678 | 8.71% | 23.49% | 32.85% | 24.81% | 0.01% |
| Week 16 | 485632 | 38358 | 8.42% | 25.06% | 27.54% | 30.70% | 0.00% |
| Week 17 | 468694 | 40899 | 7.65% | 25.46% | 27.22% | 32.85% | 0.00% |
| Week 18 | 490175 | 35863 | 8.15% | 25.23% | 29.17% | 33.01% | 0.02% |
| Week 19 | 471590 | 39960 | 7.37% | 24.30% | 31.36% | 35.25% | 0.00% |
| Week 20 | 481472 | 34757 | 5.92% | 23.44% | 31.49% | 35.88% | 0.00% |
| Week 21 | 519240 | 28503 | 4.98% | 21.38% | 30.57% | 38.74% | 0.03% |
| Week 22 | 585687 | 25840 | 4.29% | 17.70% | 31.76% | 41.28% | 0.10% |
| Week 23 | 630164 | 25137 | 4.16% | 17.27% | 39.34% | 37.76% | 0.00% |
| Week 24 | 741860 | 26222 | 3.61% | 18.68% | 29.34% | 46.40% | 0.08% |
| Week 25 | 759708 | 26773 | 3.97% | 12.94% | 21.11% | 61.80% | 0.02% |
| Week 26 | 884719 | 30174 | 3.39% | 15.35% | 29.38% | 51.97% | 0.05% |
| Week 27 | 982742 | 30020 | 2.88% | 19.80% | 34.25% | 43.44% | 0.05% |
| Week 28 | 1103844 | 28291 | 2.65% | 13.83% | 27.77% | 56.64% | 0.00% |
| Week 29 | 1224975 | 29277 | 2.52% | 17.23% | 20.04% | 60.67% | 0.08% |
| Week 30 | 1274869 | 30929 | 2.85% | 11.71% | 15.17% | 71.37% | 0.10% |
| Week 31 | 1436783 | 36325 | 2.38% | 10.42% | 12.50% | 74.69% | 0.60% |
| Week 32 | 1546074 | 34253 | 2.30% | 12.94% | 14.37% | 70.84% | 1.59% |
| Week 33 | 1543530 | 35582 | 2.53% | 13.34% | 18.03% | 64.37% | 3.87% |
| Week 34 | 1821055 | 39100 | 2.15% | 14.92% | 18.72% | 60.28% | 4.85% |
| Week 35 | 1742152 | 39240 | 2.19% | 13.31% | 22.38% | 53.13% | 9.85% |
| Week 36 | 1782811 | 38088 | 2.11% | 13.87% | 21.14% | 47.69% | 16.57% |
| Week 37 | 1886643 | 37559 | 2.15% | 14.06% | 23.92% | 42.11% | 19.16% |
| Week 38 | 1844959 | 40656 | 1.99% | 11.49% | 20.69% | 39.70% | 27.66% |
| Week 39 | 1998897 | 36764 | 1.82% | 13.93% | 18.82% | 32.39% | 34.61% |
| Week 40 | 2006967 | 36475 | 1.95% | 13.15% | 14.98% | 31.32% | 40.52% |
| Week 41 | 2026073 | 39079 | 1.94% | 12.71% | 17.31% | 29.23% | 40.54% |
| Week 42 | 2268892 | 39228 | 1.61% | 10.86% | 14.22% | 24.24% | 50.41% |
| Week 43 | 2443594 | 36547 | 1.63% | 12.01% | 13.97% | 22.69% | 51.27% |
| Week 44 | 2884604 | 39712 | 1.56% | 11.02% | 14.49% | 19.53% | 54.93% |
| Week 45 | 3355265 | 45051 | 1.63% | 14.33% | 14.65% | 16.52% | 54.44% |
| Week 46 | 3690495 | 54835 | 1.62% | 11.62% | 14.47% | 17.17% | 56.64% |
| Week 47 | 3977223 | 59699 | 1.69% | 11.32% | 12.95% | 14.36% | 61.30% |
| Week 48 | 4060891 | 67221 | 1.72% | 12.26% | 16.25% | 14.74% | 56.66% |
| Week 49 | 3935330 | 69916 | 1.87% | 15.96% | 17.86% | 13.08% | 52.94% |
| Week 50 | 3970427 | 73396 | 1.89% | 17.47% | 17.73% | 11.11% | 52.93% |
| Week 51 | 4329927 | 75038 | 1.82% | 11.99% | 12.73% | 10.05% | 65.14% |
| Week 52 | 4612790 | 79001 | 1.58% | 18.86% | 17.73% | 10.08% | 53.07% |
| Week 53 | 4068632 | 72761 | 1.87% | 24.45% | 14.08% | 8.07% | 53.40% |
| Week 54 | 4035226 | 76017 | 2.12% | 20.48% | 41.64% | 11.60% | 26.17% |

**Table S2: aa change in N protein SR rich region where the frequency is >300.**

| WildtypeAA | Position↑ | MutatedAA | Frequency↑ |
|---|---|---|---|
| S | 194 | L | 1401 |
| S | 197 | L | 1001 |
| S | 188 | L | 822 |
| S | 193 | I | 583 |
| S | 202 | N | 310 |
